# Pre-school childcare and inequalities in child development

**DOI:** 10.1101/2020.10.05.20206946

**Authors:** Michael J Green, Anna Pearce, Alison Parkes, Elaine Robertson, S Vittal Katikireddi

**Author notes:** Corresponding Author: Michael J Green.

## Abstract

Centre-based childcare may benefit pre-school children and alleviate inequalities in early childhood development, but evidence on socio-emotional and physical health outcomes is limited. Data were from the UK Millennium Cohort Study (n=14,376). Inverse-probability weighting was used to estimate confounder-adjusted population-average effects of centre and non-centre-based childcare (compared to parental care only) between ages 26-31 months on (age 3): internalising and externalising symptoms, pro-social behaviour, independence, emotional dysregulation, vocabulary, school readiness, and body mass index. To assess impacts on inequalities, controlled direct effects of low parental education and lone parenthood on all outcomes were estimated under two hypothetical scenarios: 1) universal take-up of centre-based childcare; and 2) parental care only. On average, non-centre based childcare improved vocabulary and centre-based care improved school readiness, with little evidence of other benefits. However, socio-economic inequalities were observed for all outcomes and were attenuated in scenario 1 (universal take-up). For example, inequalities in externalising symptoms (according to low parental education) were reduced from a confounder-adjusted standard deviation difference of 7.8 (95% confidence intervals: 6.7, 8.8), to 1.7 (0.6, 2.7). Inequalities by parental education in scenario 2 (parental care only) were wider than in scenario 1 for externalising symptoms (at 3.4 (2.4, 4.4)), and for emotional dysregulation and school readiness. Inequalities by lone parenthood, which were smaller, fell in scenario 1, and fell further in scenario 2. Universal access to centre-based pre-school care may alleviate inequalities, while restricted access (e.g. during lockdown for a pandemic such as Covid-19) may widen some inequalities in socioemotional and cognitive development.

## Introduction

Children from less socio-economically advantaged families tend to experience worse health and poorer socio-emotional and cognitive development than their more advantaged peers [1, 2]. Inequalities may arise via many pathways, including material deprivation and disadvantages in parental psychosocial resources, which can have negative impacts on parenting capacity [3-5]. Investment in early years is widely accepted as one of the most effective ways to reduce inequalities in childhood and across the life course [6-8] and childcare and early years’ learning can be important domains of governmental policy. Early childhood education and care (ECEC) may benefit children’s cognitive and social development [9-11], though a recent meta-analysis of natural experiment studies found mixed evidence, with the most consistent positive effects for cognitive and academic outcomes, higher-quality programmes, and publicly-funded provision [12]. Non-centre childcare by grandparents has also been shown to support language development [9] and emotional wellbeing [13]. Mechanisms of benefit may include provision of cognitive or academic training and social experiences with other children and adults, boosting confidence and easing transitions into school-based settings [9].

Provision of universal, cost-free and high-quality childcare has potential to reduce inequalities in children’s outcomes, giving all children a stable, nurturing and educationally stimulating setting. Several studies have shown that centre-based childcare yields greater cognitive and academic benefits for children from socio-economically disadvantaged families [9, 12, 14, 11]. Evidence in relation to socio-emotional outcomes is more mixed. Centre-based childcare may buffer against risk factors that are more prevalent among disadvantaged families. For example, childcare may reduce effects of maternal depression on children’s internalising symptoms [15]. However, childcare has also been associated with greater socio-emotional benefits for children from more advantaged families [16], which is perhaps due to inequalities in the quality of care families receive. Differential benefits of non-centre care are also uncertain. Non-centre care by grandparents can buffer against disadvantage [17-19], and can be especially beneficial where there is only one resident parent [20], but has also been shown to exacerbate socioeconomic inequalities in cognitive development [14].

Evidence for impacts of childcare on physical health is less well developed and more varied. For example, group childcare can increase risk of infectious disease [11, 21], and there is mixed evidence of impacts on children being overweight or obese. Some, but not all, studies show decreased risk of being overweight associated with centre-based care. Again, the quality of care provision may be important, and non-centre care (e.g. with grandparents or other relatives/friends) may be associated with increased risk [10, 22-24, 13, 25, 26]. Furthermore, there is little longitudinal evidence for effects of childcare on diet and physical activity [27], and some studies have shown that childcare, especially non-centre care, was more strongly associated with children being overweight in more advantaged households [13, 28-30].

ECEC is a key feature in government policies across the globe, and many high-income countries have large proportions of children experiencing some ECEC. For example, in the UK ECEC has been offered universally and free of charge to all 3-4 year olds since 2004. This ECEC entitlement has expanded since its introduction and by 2012 ranged from 15-30 hours per week. Free childcare hours for children aged 2 years in the UK is still means-assessed and in early 2020 was taken up by 69% of those eligible [31]. For many working parents, non-centre-based childcare (e.g. with family, friends, childminders etc) remains crucial, to cover gaps between what is provided and what is needed. Yet despite the increased provision of ECEC, we have yet to establish its potential to alleviate inequalities in health and development. The question of whether children benefit differentially from ECEC has become particularly relevant during the COVID-19 pandemic. Many countries, including the UK, have enacted social mitigation ‘lockdown’ measures, reducing physical proximity to others to slow infection transmission [32-35]. This has meant temporary withdrawal of almost all centre-based childcare provision (excepting children of key workers, who may still be eligible for centre-based care). Non-centre-based childcare, by family and friends from separate households, will also have been largely impossible (though perhaps not needed if working parents were furloughed). This could widen inequalities in cognitive and socio-emotional development if children from disadvantaged households derive more benefit from childcare than those from more advantaged households [33, 34]. Impacts could also extend to other outcomes, as, for example, less advantaged families may find it harder to provide healthy nutrition without the meals provided with ECEC childcare [34, 35].

Historic data with a mixed distribution of those using parental, non-centre and centre-based childcare and may yield useful estimates in relation to the following questions, for a range of relevant child outcomes. Question 1 aims to anticipate the average effects of lockdown restrictions on child outcomes. Question 2 seeks to understand how expanded access to centre-based childcare could have impact on inequalities in child outcomes, while question 3 addresses how social mitigation measures removing access to both centre and non-centre-based childcare may impact inequalities:

1. What are effects of restricting access to centre and non-centre-based pre-school childcare for a period of 6 months in early life?
2. What impact might universal take-up of centre-based pre-school childcare have on inequalities by parental education and family structure?
3. What impact could universal restriction of access to centre and non-centre-based pre-school childcare have on inequalities by parental education and family structure?

We address question 1 by estimating average effects of centre and non-centre-based childcare (as opposed to parental care) on child outcomes using inverse probability weighting. We address questions 2 and 3 using mediation analyses to estimate effects of education and family structure on child outcomes under two hypothetical scenarios; one where all children receive centre-based care (question 2), and one where all children receive parental care only (question 3).

## Methods

### Data

Data were from the UK Millennium Cohort Study (MCS), a nationally representative survey of children born in the UK, September 2000-January 2002 [36]. A stratified clustered sampling design was used to oversample children living in Wales, Scotland and Northern Ireland, disadvantaged areas and, in England, areas with high proportions of ethnic minority groups. Families were selected through Child Benefit Records, and initially contacted for opt-out by the Department for Work and Pensions. Initial interviews took place at 9 months, when information was collected on 72% of those contacted, providing information for 18,818 children (18,296 of whom were singletons). Follow-up interviews were conducted at approximately age 3 years (76.4% of the original sample; mean age of child was 38.2 months). Data were analysed for 14,376 singleton children whose mothers were interviewed at baseline and follow-up. Partners were also interviewed where applicable and if possible.

### Measures

#### Parental Education and Family Structure (Primary Exposures)

Parents reported their highest educational qualifications at baseline (9 months) and these were coded to compare either parent having A-Level qualifications (or higher) against parents with lower or no qualifications (i.e. differentiating between those who had and had not completed qualifications post-compulsory education). Baseline family structure was coded to compare single parent households against two-parent households (natural parent couples or reconstituted families).

#### Pre-school Childcare (Mediator)

Mothers were asked about their main childcare arrangement at baseline (9 months) and if and when this had changed at the age 3 interview. Childcare type was classified as “parental” if the child was only cared for by the mother, father or the mother’s partner; “non-centre-based” if they were also cared for by a friend, neighbour, grandparent, other relative, babysitter, childminder, nanny or au pair; and “centre-based” if they were cared for in a nursery, play group or childcare centre. Mothers were also asked at both interviews if they had regularly used any other form of child-care, and if so when that had started and ended. If the primary childcare arrangement was “parental” but an additional arrangement involved non-parental childcare, then this additional childcare type was used in order to assess any regular exposure to non-parental childcare. The three categories used for analysis were: parental care only; some non-centre-based care (but no centre-based care); and some centre-based care (potentially in combination with some non-centre-based care).

At the age 3 interview, mothers were asked how long centre and non-centre-based childcare arrangements had been in place. However, the child’s age at this interview varied somewhat, with the youngest age being 32 months. In order to standardise the period of childcare assessment across all participants, the childcare variable was coded to represent the specific 6-month period where the child was aged 26-31 months. We also conducted supplementary analyses varying the exposure period to 3 months (ages 29-31 months) and 12 months (ages 20-31 months).

#### Socio-emotional wellbeing, cognitive development and body mass index (child outcomes)

Child outcomes were all assessed at the age 3 survey. Mothers completed the Strengths and Difficulties Questionnaire [37-39]. The emotional symptoms and peer problems dimensions were combined into an internalising symptom score, and the conduct problems and hyperactivity/inattention dimensions were combined into an externalising symptom score [40]. The pro-social dimension was considered as a separate outcome. Two sub-scales from the Child Social Behaviour Questionnaire were also completed by the mother and respectively assessed independence and self-regulation (e.g. works things out for self, persists with difficult tasks) and emotional dysregulation (e.g. easily frustrated, shows mood swings) [37]. Cognitive tests were administered by trained researchers. The naming vocabulary sub-test from the British Ability Scales II [41] involves the child being shown pictures of objects and asked to name them. Scores were standardised for child age (in months) at time of interview [42], with higher scores indicating a more expansive vocabulary. The Bracken School Readiness Assessment-Revised (BSRA-R) [43] was also completed and assesses knowledge of basic concepts such as numbers, letters, shapes and colours. Scores were again standardised for age at interview [42]. Children’s weight and height were measured without shoes or outdoor clothing by trained interviewers, using Tanita HD-305 scales (Tanita UK Ltd, Middlesex, UK) and the Leicester Height Measure Stadiometer (Seca Ltd, Birmingham, UK). Body mass index (BMI, kg/m^2^) z-scores were calculated using the “zanthro” command in Stata [44]. Estimated effects are shown in standard deviation units for all outcomes to facilitate comparison.

#### Confounders

We adjusted for a range of potential confounders relating to socio-demographic factors, parental health and health-related behaviours, and parenting style. In order to avoid deterministic relationships with our family structure measure, where characteristics were measured on both parents we took a family level approach (i.e. using either parent’s status for couples, whichever was least optimal).

Child sex; ethnicity (White UK vs ethnic minority); UK country (England, Scotland, Wales or Northern Ireland); mother’s age at first live birth (<19, 20-24, 25+); and child temperament, measured with the Carey Infant Temperament Scale [37], were all captured at 9 months. The following were measured at both age 9 months and 3 years: poverty (indicated by equivalised household income <60% of the median); housing tenure (owned/mortgage vs rented/other); parental economic activity (at least one parent in any kind of paid employment); parental occupational class (3 category coding from the UK National Statistics Socioeconomic Classification for last known job, taking the more advantaged class from couple parents); parental mental health (at baseline using a modified 9-item version of the Rutter Malaise Inventory [45, 37], at age 3 the 6-item Kessler scale [37, 46]); parental drinking frequency (either parent consuming alcohol >5 times a week vs less frequently); parental smoking (either parent currently smokes vs neither parent currently smokes); and whether either parent had a longstanding illness (baseline but not age 3 questions further differentiated longstanding illnesses that were perceived as limiting). Measures from the age 3 survey included: the Pianta parent-child relationship conflict and warmth scales (taking the higher score from either resident parent for conflict and the lower score for warmth) [37, 47]; Straus’ conflict tactics scale as an indication of negative discipline (mother reported) [48, 37]; home ‘disorganisation’ measured with 3 items from the confusion, hubbub and order scale (mother reported) [49]; a measure of home ‘routine’, based on two items asking about the extent to which the child has regular bedtimes and mealtimes (mother reported) [37]; a measure of parental involvement in educational activities at home (mother reported) [50]; an interviewer assessment of how emotionally supportive the home environment was [37, 51] and presence of a younger sibling, other siblings (0, 1, 2 or more), or other adults in the household.

We also created a variable indicating experience of centre or non-centre-based childcare at earlier ages (<26 months) and included a later measure of family structure (lone parent vs couple at age 3). Adjustment strategies for how these variables were included in our analyses are discussed below.

#### Analytic sample

All analyses were weighted for sampling and drop out at the age 3 follow-up. Table 1 shows proportions of the sample who had missing data on each variable. While only 9,077 (63.1%) had full data on all variables, another 2,695 (18.8%) were missing data on a single variable, and 2,604 (18.1%) were missing data on more than one of the analysis variables. We performed multiple imputation (25 datasets) in order to include all 14,376 respondents at follow-up, reduce effects of differential response bias, and maximise use of observed data.

**Table 1:**
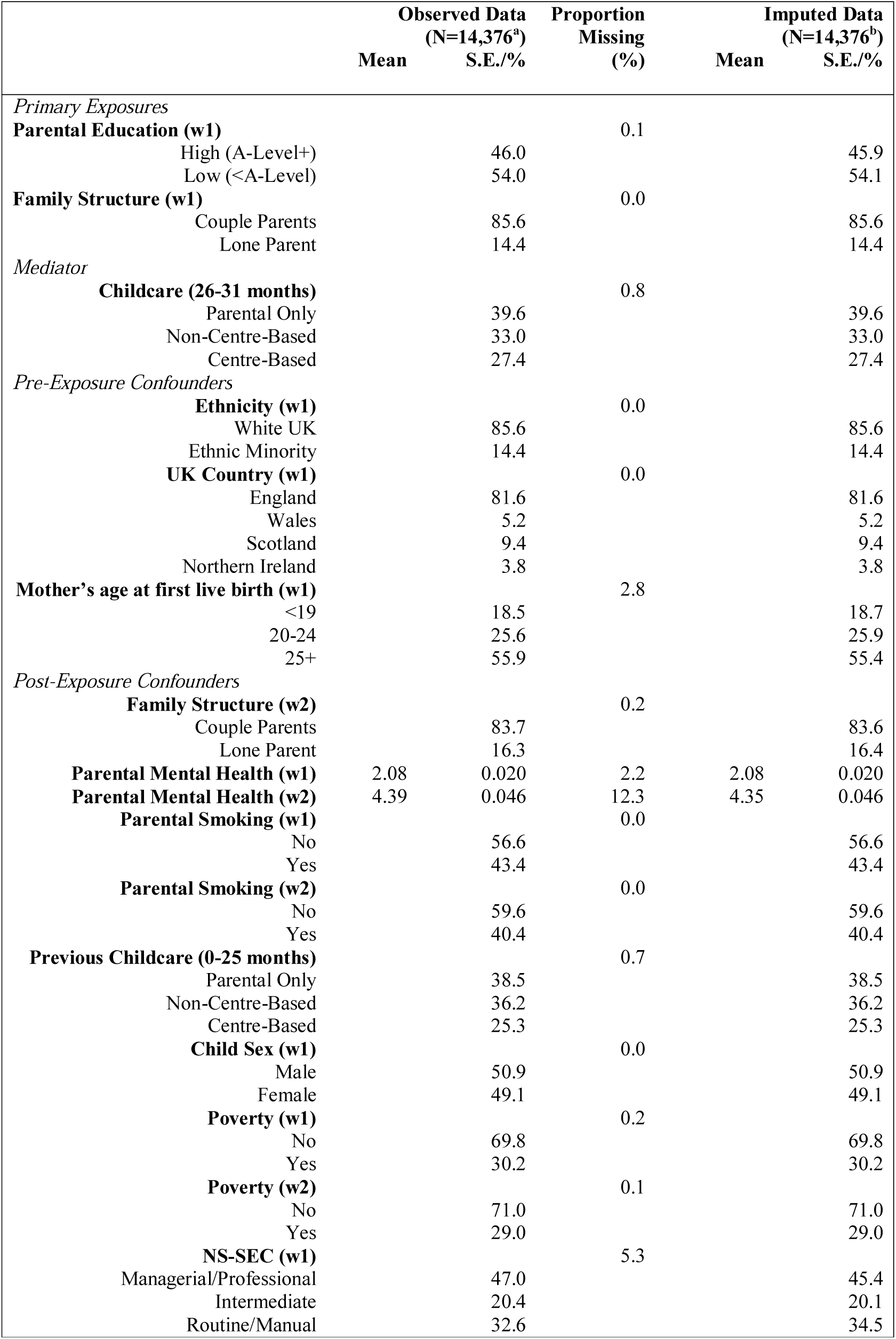
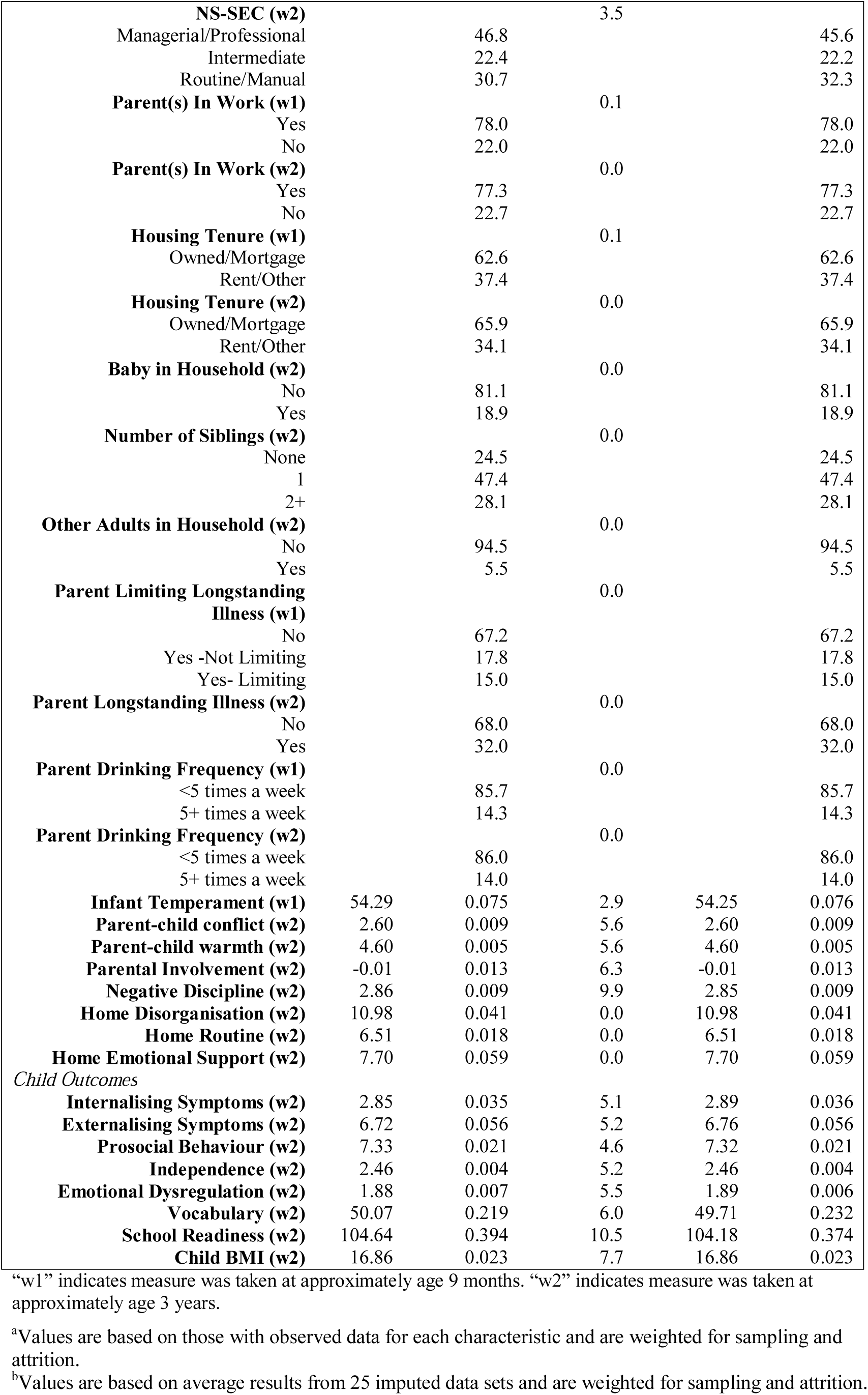
Descriptive Statistics

#### Approach to confounding

To address Question 1, we examined the association between childcare and the children’s outcomes adjusting for all the confounders listed above as well as for education and family structure (which were viewed as confounders for the average effects of childcare). For Questions 2 and 3, we assume that pre-school childcare mediates effects of either parental education or family structure on child and parent outcomes, as shown in Figure 1. We make a distinction between pre-exposure confounders (C) and post-exposure confounders (L). Pre-exposure confounders are potential common causes of the exposure, the mediator and the outcome. In contrast, post-exposure confounders are potential common causes of the mediator and the outcome but may (or may not) be caused by the exposure. This distinction is important for estimating the effect of the exposure after intervention on the mediator [52]. Table 2 shows which variables were considered as pre/post confounders. The causal direction of relationships between childcare and age 3 measures was ambiguous for many of the post-exposure confounders. Our main results assume these confounders are determinants of childcare, but we conducted sensitivity analyses where the age 3 measures were assumed to be caused by childcare (and therefore not adjusted).

**Table 2:**
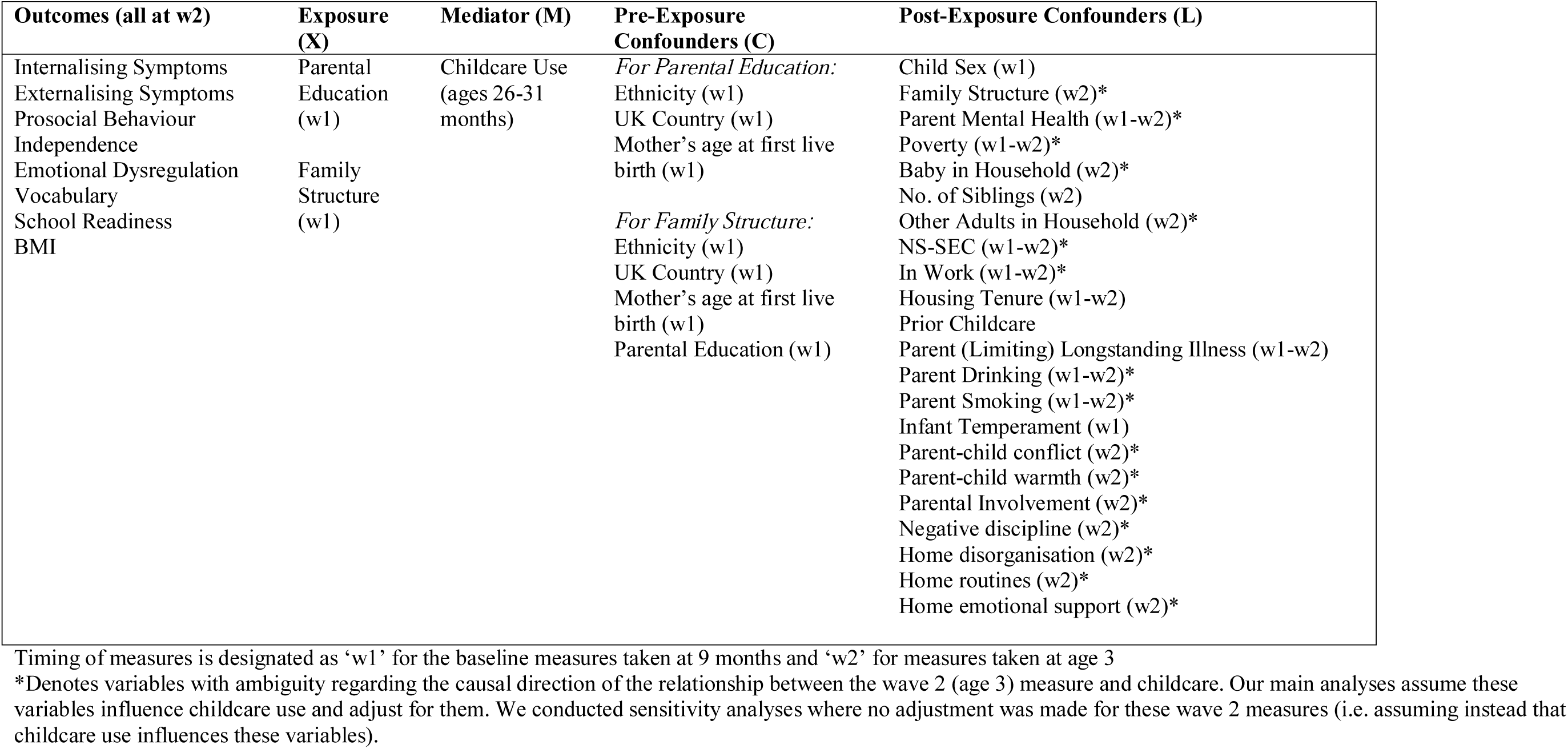
Analysis Variables

**Figure 1:**
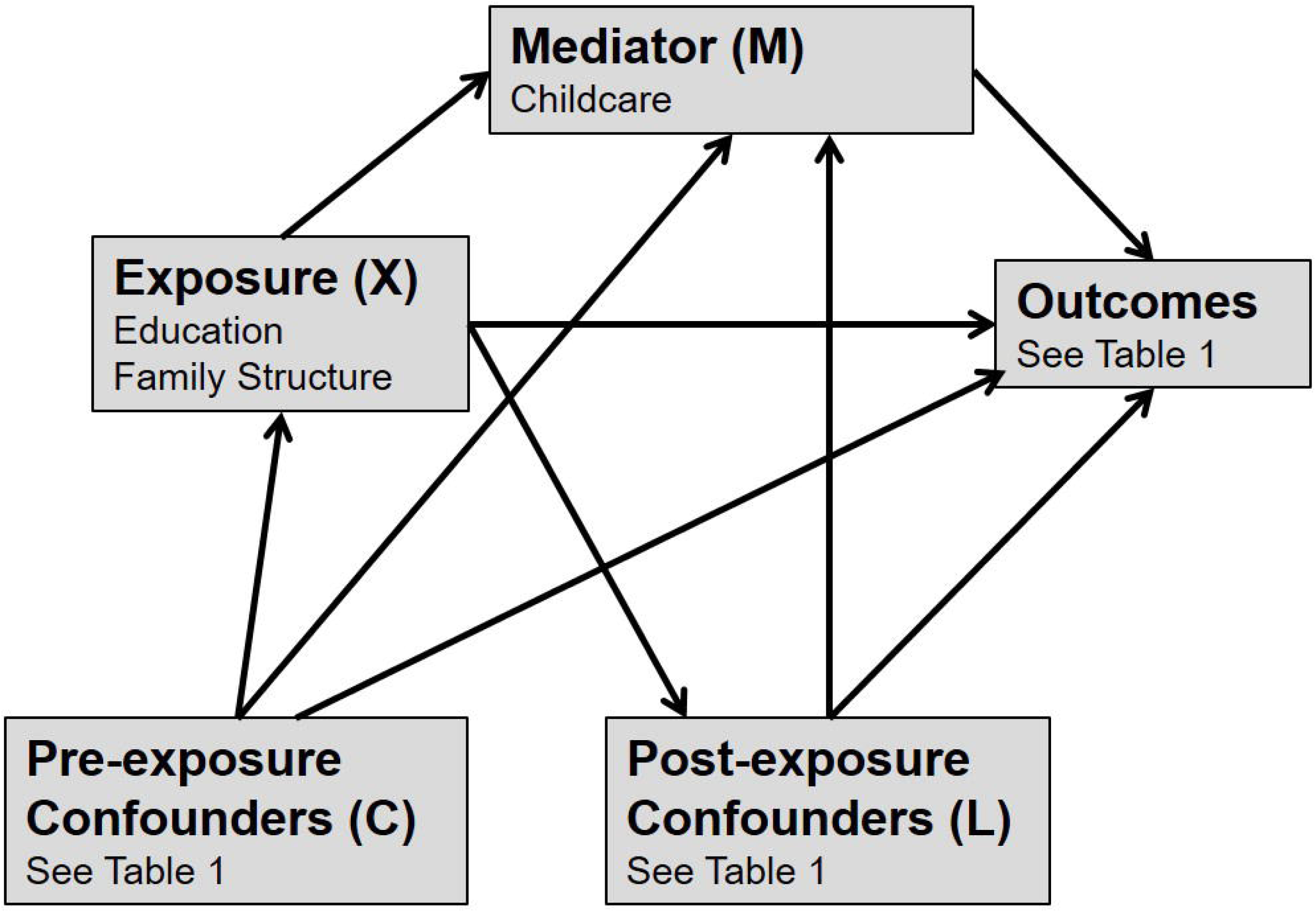
Assumptions about causal direction in our analyses

#### Analysis for Question 1: Estimating Childcare Effects

We first estimated effects of six months’ exposure to centre and non-centre-based pre-school childcare on all outcomes, using an inverse probability weighting (IPW) procedure (described more fully in Supplementary File 1) to adjust for all confounders and for parental education and family structure. Estimates represent Average Treatment Effects (ATEs) [53], i.e. the average effects of pre-school childcare within the sample. Supplementary File 1 contains more details on calculation of the weights and shows that they balanced observed confounders between childcare categories.

#### Analysis for Questions 2 and 3: Estimating Impacts on Inequalities

Total effects of parental education and family structure were estimated using a similar IPW approach to adjust for pre-exposure confounders. Post-exposure confounders were not adjusted for here because effects via these post-exposure variables were part of the total effect we were trying to estimate. We again estimated ATEs, i.e. the average effect of parental education or family structure within the sample. Supplementary File 1 contains more details on calculation of the weights and shows that they balanced observed pre-exposure confounders between exposure categories.

Finally, we estimated controlled direct effects (CDEs) of each exposure on all outcomes. The CDE represents the estimated effect of the exposure under hypothetical intervention to set the mediator (childcare) to the same value for everyone [52]. CDE estimation explicitly allows for interaction between the exposure and mediator in their effect on the outcome, such that CDEs can differ depending on the level of childcare respondents are all set to receive. We estimated two CDEs for each outcome. The first scenario set all respondents to receive some centre-based childcare, representing potential impacts on inequalities if universal take-up were achieved. This may also approximate the UK pre-pandemic context, where take-up was already high [31]. The second scenario, with childcare set to parental care only for all respondents, approximates what may have happened under lockdown restrictions. CDE estimates were derived from inverse-probability weighted marginal structural models [52] and adjusted as above for pre-exposure confounders. However, in contrast to traditional regression methods, this allows adjustment for differences in post-exposure confounders that are *not* due to the exposure, without removing differences that *are* due to the exposure (and therefore part of the desired effect). Thus, the path from exposure (X) to the outcome via confounders (L) in figure 1 remains open and is included in the CDE estimate. Supplementary File 1 contains details on calculation and performance of these weights. While the weights mostly balanced confounders as expected, there was some residual imbalance among lone parents, suggested that data were insufficient to fully disentangle differences in childcare use within this group from differences in observed confounding factors (though confounders were balanced among couple parents, i.e. the majority of the sample).

## Results

### Sample Description

Table 1 shows descriptive statistics for the sample, and compares the observed and the imputed data, showing little difference in sample characteristics between the two. Over the 6 months from 26-31 months of age, 27.4% of the sample had used centre-based childcare, while 33.0% had used only non-centre-based childcare.

### Estimating Effects of Childcare on Child Outcomes

Figure 2 shows unadjusted and adjusted associations between centre and non-centre-based childcare (as compared to parental care only) and each outcome. Whilst many of the outcomes were associated with use of childcare, the confounder-adjusted ATE estimates only indicated the following clear differences: those using non-centre-based care had higher vocabulary scores; and those using centre-based care had higher school readiness scores.

**Figure 2:**
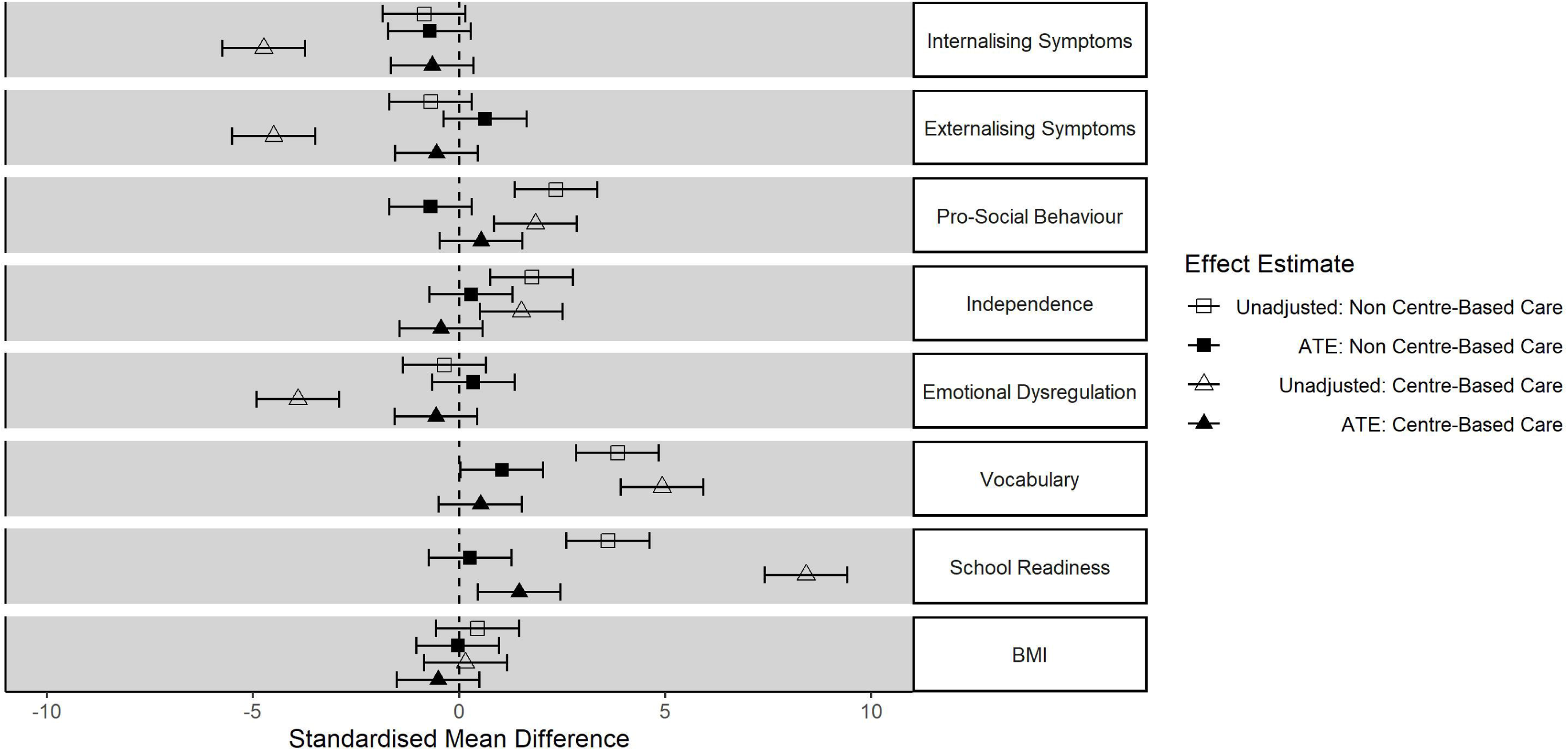
Estimated effects (and 95% confidence intervals) of centre and non-centre-based childcare on child outcomes compared to parental care only (from ages 26-31 months)

### Inequalities by Parental Education

Figure 3 shows inequalities in child outcomes by parental education. The confounder-adjusted ATE estimates imply that, in the observed data, there are strong inequalities. Low parental education was associated with more externalising and internalising symptoms, less pro-social behaviour and independence, more emotional dysregulation, and lower vocabulary and school readiness. Child BMI was the only outcome for which parental education did not show a clear effect.

**Figure 3:**
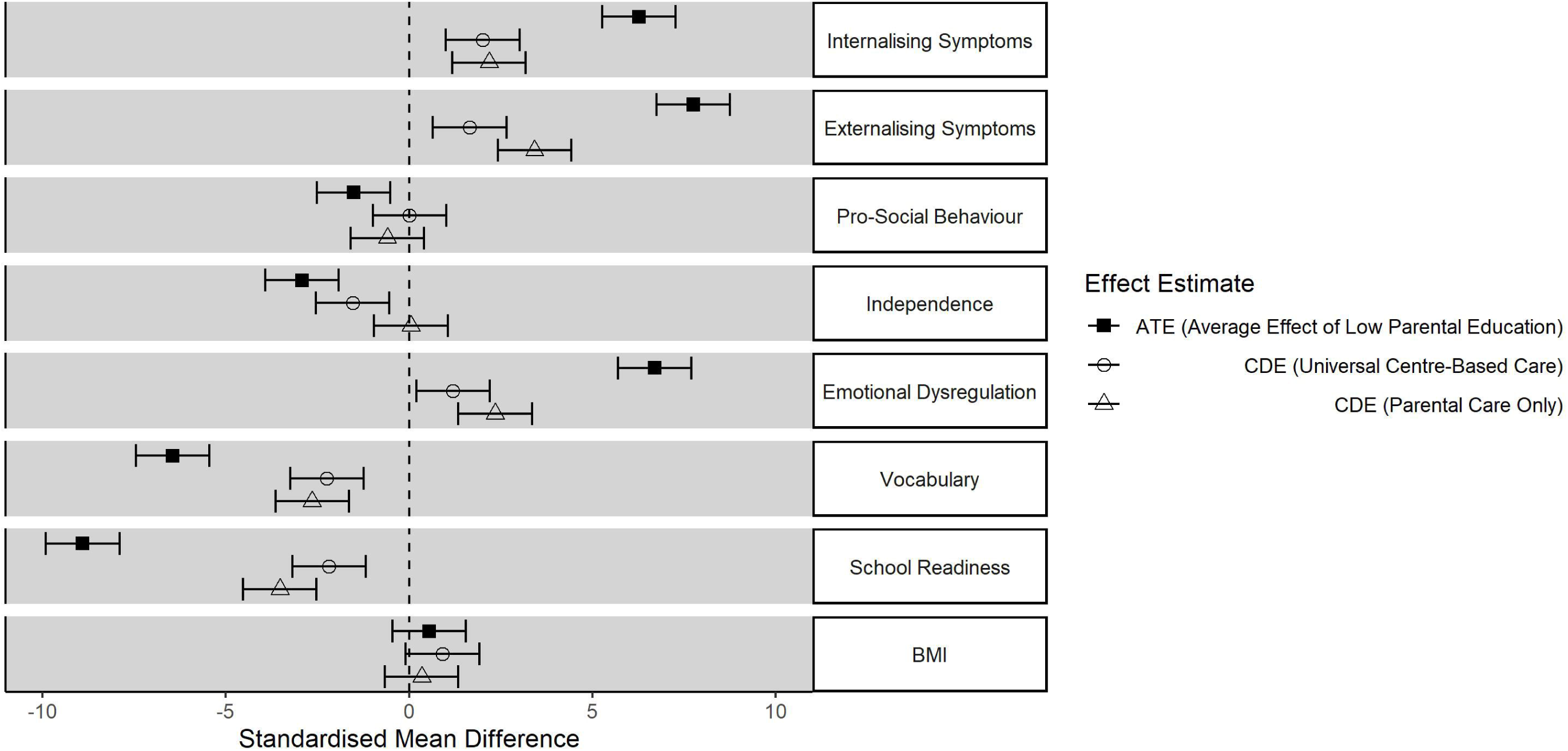
Inequalities (and 95% confidence intervals) in child outcomes by parental education (Low vs. High), in the observed data (ATE), scenario 1 (universal centre-based care) and scenario 2 (parental care only)

Inequalities were substantially attenuated in CDE estimates after simulating universal use of centre-based care while the child was aged 26-31 months (scenario 1). In scenario 2 (universal restriction to parental care) inequalities were also attenuated although not to the same extent as seen in the universal centre-based care scenario. Specifically, scenario 2 showed wider inequalities in externalising symptoms, emotional dysregulation and school readiness than in scenario 1. Given that the observed inequalities are based on historical data and that scenario 1 probably better reflects the childcare situation in the UK before the COVID-19 pandemic, this would imply that a move to parental care only could increase inequalities.

### Inequalities by Family Structure

Figure 4 shows inequalities in child outcomes by baseline family structure. Inequalities were smaller in magnitude than those associated with parental education, but there were still clear effects including more externalising and internalising symptoms, more pro-social behaviour, lower vocabulary and school readiness, and higher BMI among children living in single parent households.

**Figure 4:**
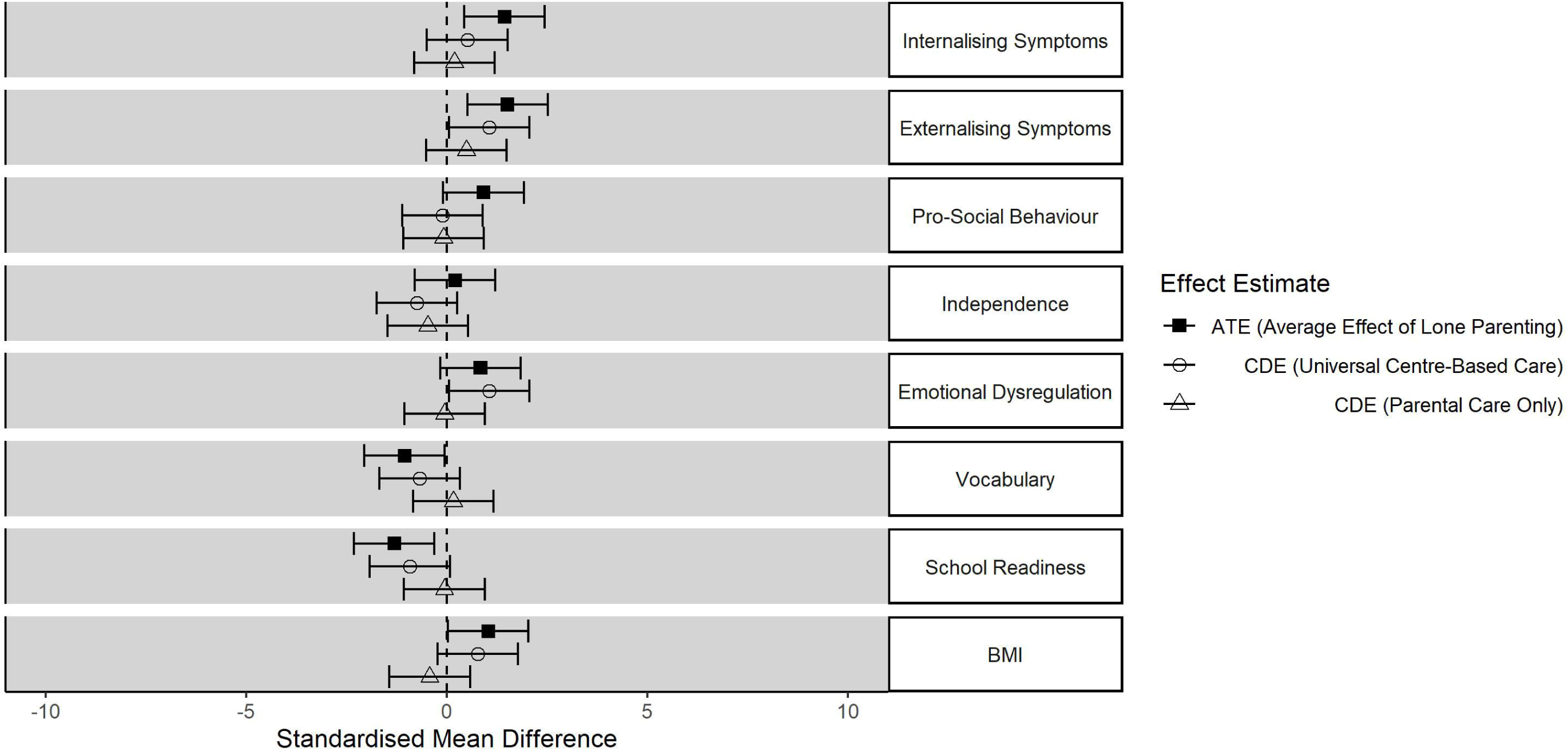
Inequalities (and 95% confidence intervals) in child outcomes according to lone parenthood, in the observed data (ATE), scenario 1 (universal centre-based care) and scenario 2 (parental care only)

These inequalities were somewhat attenuated in scenario 1 (universal take-up of centre-based care), although clear inequalities in internalising symptoms and school readiness remained. In contrast to the analyses relating to parental education, universal restriction to parental childcare (scenario 2) produced further attenuation of these inequalities.

### Sensitivity Analyses

Supplementary File 2 repeats the main analyses with childcare defined over periods of 3 and 12 instead of 6 months (respectively covering ages 29-31 months and 20-31 months), and with a more restricted set of post-exposure confounders included (as indicated in Table 1). Findings were similar.

## Discussion

With observational data from this large and nationally representative sample of UK children born at the turn of the century, we estimated that, compared to parental care only between the ages of 26-31 months, centre and non-centre-based childcare were associated with some improvements in school readiness and vocabulary respectively. This was consistent with findings from natural experiment studies [12], where the strongest evidence has been for effects on cognitive/academic outcomes. While these average effects were relatively minor, they may mask differential effects, and we estimated considerable impacts of childcare on inequalities for a range of socioemotional, cognitive and physical outcomes. Compared to inequalities in the observed data (in which one fifth of children attended centre-based childcare and one third attended non-centre based care), inequalities by parental education and family structure were considerably attenuated in a scenario simulating universal use of centre-based care. This indicates the potential impact of achieving universal take-up of centre-based care at these ages, but relative to the observed data also probably more closely reflects conditions for UK children before the covid-19 pandemic, where uptake of childcare was already high [31]. In a scenario simulating universal restriction to only parental care (approximating covid-19 lockdown impacts) inequalities by parental education were also reduced (as compared to the observed data) but remained considerably larger than those in scenario 1 for externalising symptoms, emotional dysregulation and school readiness. This suggests that children of less educated parents may derive more benefit from centre-based care than those with more educated parents, as other studies have indicated [12, 11, 9]. Inequalities related to lone parenthood, while smaller, were actually further attenuated in this second parental-care-only scenario, suggesting that children born in couple parent families were deriving greater benefits from centre-based care than those born in lone parent families.

Our findings suggest that policies extending access to centre-based childcare for pre-school children may alleviate inequalities in children’s socioemotional, cognitive and physical well-being. Measures (such as lockdowns) that restrict access to centre and non-centre-based childcare in contexts where uptake was otherwise high may exacerbate inequalities by parental education in some socioemotional and cognitive outcomes. Re-establishing childcare access should be an important goal amidst necessary pandemic mitigation measures. The further attenuation of inequalities by family structure in the parental care only scenario, likely represents removal of privilege among advantaged groups rather than a levelling up of inequalities. This may be because couple families have access to better quality childcare, or because the stresses involved in utilising childcare (such as juggling drop-off and pick-up times with work and other schedules, or arranging wrap-around care) are greater among lone parents, meaning they derive fewer benefits. Future research (qualitative and quantitative) is required to explore these potential pathways and highlight how the needs of children from different family structures can be better supported through childcare services.

Our findings are based on assumptions including those of no residual confounding or reverse causation. Our estimates also only represent an approximation of changes occurring in real life today. The UK expansion of access to centre-based care since 2004 may mean that centre-based care is currently very different in terms of structure, quality and accessibility today than it was when the MCS children were of a preschool age. Furthermore, universal take-up does not necessarily ensure the same quality or frequency of childcare to all families equally, and there is evidence, for example, that more hours in childcare can lead to stronger effects [12, 54]. There might also have been unmeasured inequalities in the quality of parental care: more advantaged parents may have access to better resources such as books, educational games, more indoor and outdoor space, and may feel more confident or capable in using such resources. Nevertheless, in coding our variables for *any* regular use of centre-based care, and including interactions between inequality exposures and childcare, our analyses do allow for residual inequalities in such factors.

In the context of the covid19 pandemic, which this work may inform, lockdown restrictions on childcare have not been completely universal, with some access maintained for those designated as key-workers. Furthermore, forced parental care during lockdown is not necessarily equivalent in its effect to parental care by choice or circumstance at other times and there may be inequalities in how parental care is experienced during a pandemic. While many ECECs and schools have taken concerted steps to provide materials to support parents and children, the extent to which providers have been able to do this has varied and favoured those in more advantaged circumstances [55-57]. Outcomes in our analyses were measured at approximately age 3 and it remains unclear how much any effects or any impacts on inequalities will persist as children continue to develop. There is some evidence that childcare effects can persist into later childhood, for example, childcare has been shown to be associated with reduced socioeconomic inequalities in teenage aggression [58]. Moreover, we have focused on one mechanism (childcare access), while there are a range of mechanisms related to pandemic mitigation measures which could affect children [35]. Natural experiment or time-series studies are needed to investigate the total effect of all social mitigation mechanisms on the well-being of pre-school children, including any mid to long-term effects of experiencing social mitigation at such a crucial point in the life course.

In conclusion, findings suggest average effects of centre and non-centre-based care are relatively minor, but benefits vary among socio-demographic groups. Universal take-up of centre-based childcare before age 3 may reduce socioeconomic inequalities in children’s socioemotional, cognitive and physical well-being. Where take-up has already been high (such as in the UK before the covid-19 pandemic), restrictions on childcare access may exacerbate inequalities by parental education in externalising symptoms, emotional dysregulation and school readiness, although inequalities between children of lone and couple parent families may further reduce (with children from couple parent families deriving more benefit from centre-based care). More research is needed to understand how the effects of childcare fit into those of wider contextual changes (including the covid-19 pandemic), and how to better support children from lone parent families through childcare services.

## Data Availability

Data are available from the UK Data Service.

https://beta.ukdataservice.ac.uk/datacatalogue/series/series?id=2000031

## Declarations

### Consent to Participate

This is an observational study involving secondary analysis of anonymised data from the UK Millennium Cohort Study and no ethical approval was deemed necessary. The original study obtained informed consent from parents/legal guardians of study members.

### Consent for Publication

Not applicable.

### Data/Code availability

Data are available from the UK Data Service: https://beta.ukdataservice.ac.uk/datacatalogue/series/series?id=2000031

Analytical code can be made available upon request.

### Author’s contributions

Michael Green conceived the study conception and all authors contributed to design of the analyses. Data preparation and analysis were performed by Michael Green. The first draft of the manuscript was written by Michael Green and Anna Pearce and all authors contributed to critical revisions of the manuscript. All authors read and approved the final manuscript.

