## Supplementary File 1 for "Pre-school childcare and inequalities in child development"

Article Title: **Pre-school childcare and inequalities in child development**

Journal name: **European Journal of Epidemiology**

1. MRC/CSO Social & Public Health Sciences Unit, University of Glasgow.
2. Public Health Scotland.

**Supplementary File 1**

This supplementary file contains details of the inverse probability weighting scheme applied within the main analyses.

*Estimating Effects of Childcare:*

Outcomes were regressed on childcare variables with each respondent assigned a weight equal to P(M)/P(M|X,C,L). M was their observed level of childcare within the period in question. X represents parental education and family structure which are confounders for the average treatment effects (ATE) of childcare. C and L represent sets of pre and post-exposure confounders as listed in Table 1 of the main paper (the pre/post distinction is not important here but is important for estimating impacts on inequalities). The purpose of this weighting is to balance observed confounders (X, C and L) across levels of childcare exposure (M). The probabilities required to calculate these weights were estimated via logistic regression models (with and without X, C and L). Figure S1.1 shows standardised mean differences in all these confounding variables associated with centre and non centre-based childcare, both before and after application of these inverse probability weights. Standardised mean differences within the dashed lines are those >-0.2 and <0.2 and were considered negligible. While many of the differences associated with childcare use observed using only the sampling weights were above this level the weighting reduced all differences to below this level.

Supplementary File 2 includes a re-analysis where several of the confounders (as indicated in Table 1) were assumed to be influenced by (rather than influencing) childcare use, so an alternate set of weights were used which did not include these variables. Figure S1.2 shows standardised mean differences for all confounders using this alternate (less conservative) weighting scheme. Even without including these variables in the weighting scheme, most of the differences associated with childcare were removed when weighting on the retained variables, parental economic activity at age 3 was the only variable for which there remained a large association, and that only with non centre-based childcare.

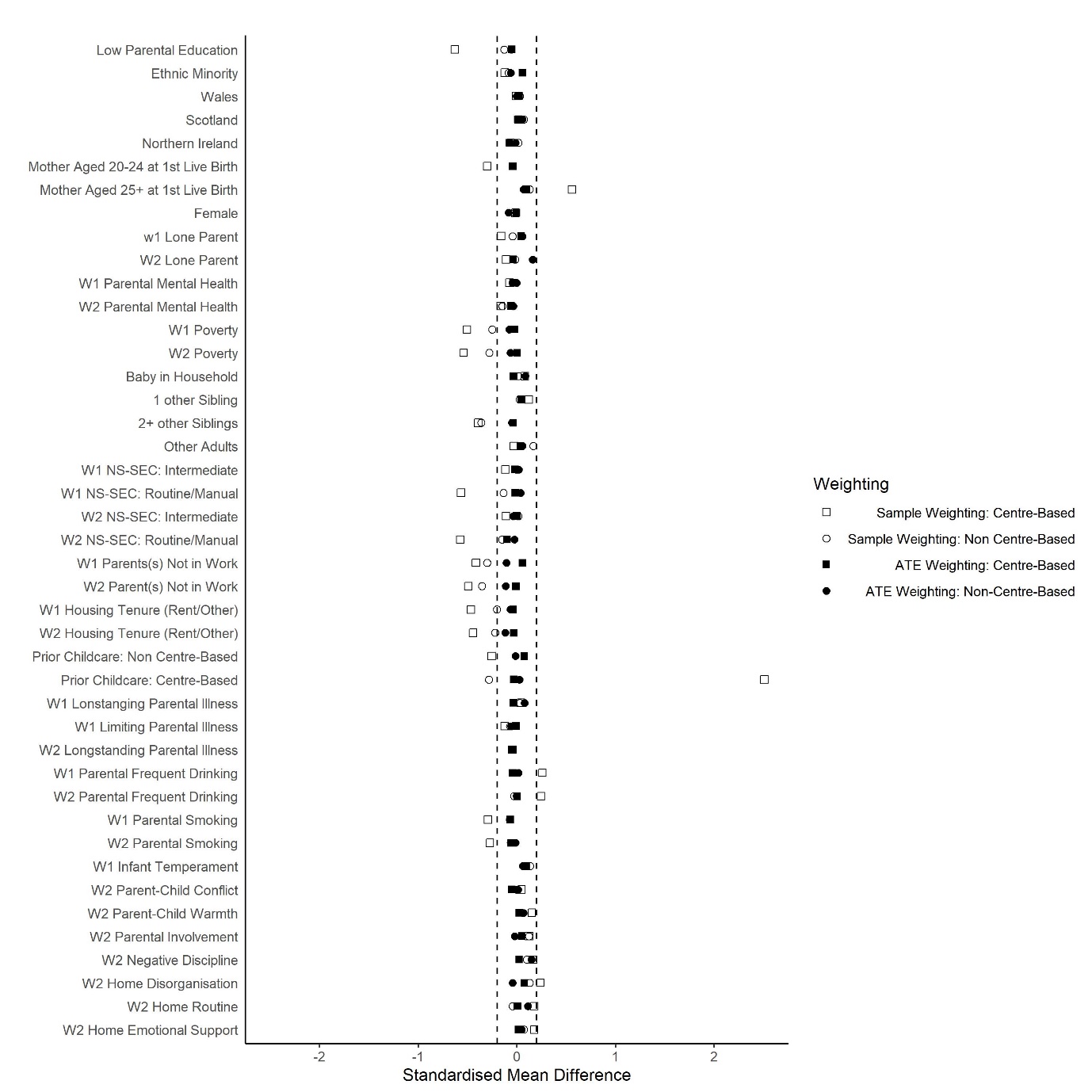

**Figure S1.1: Standardised Mean Differences in Confounders Associated with Childcare**

W1 indicates measures taken at 9 months, while W2 indicates measures taken at age 3.

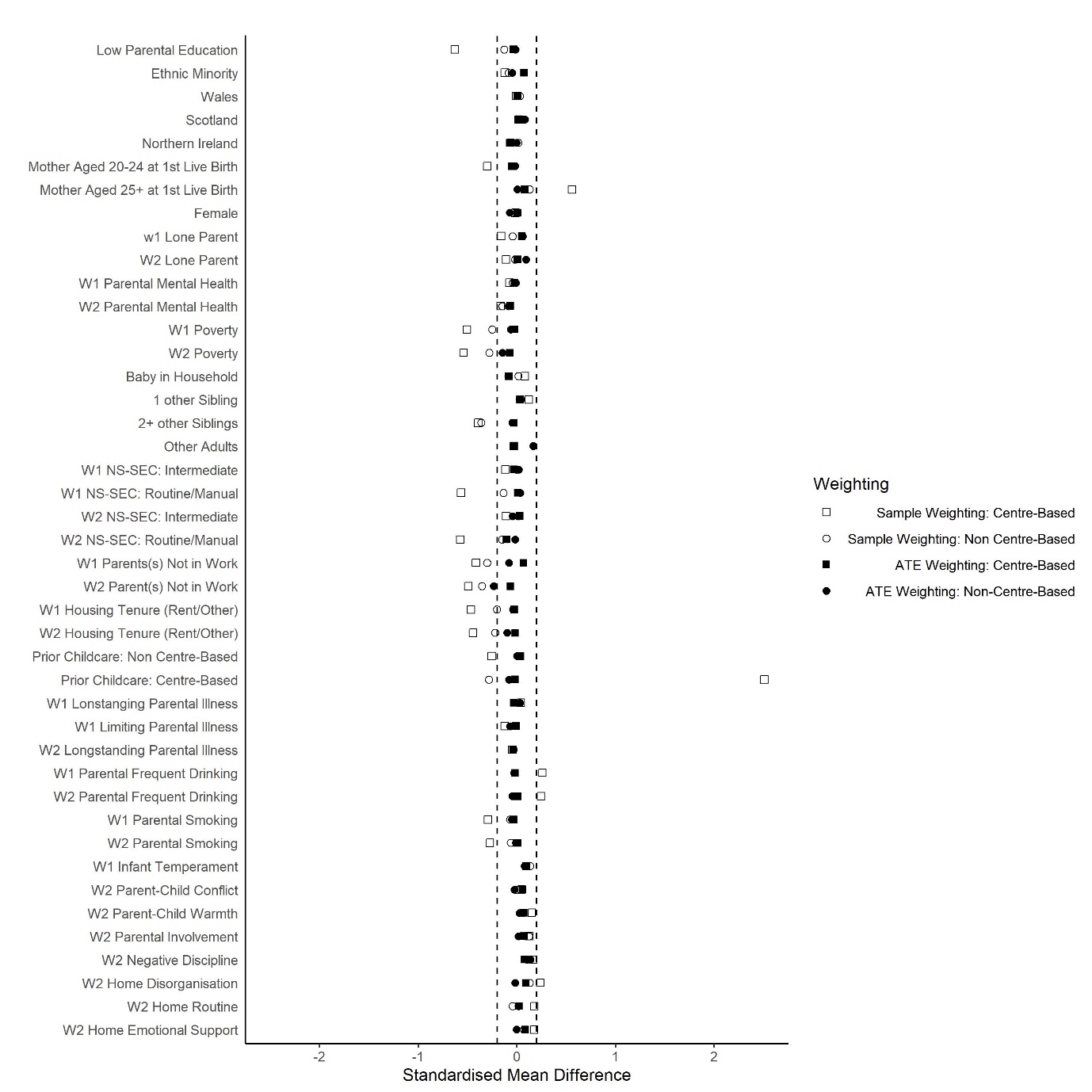

**Figure S1.2: Standardised Mean Differences in Confounders Associated with Childcare (alternate ATE weights)**

W1 indicates measures taken at 9 months, while W2 indicates measures taken at age 3.

*Estimating Average Treatment Effects (ATEs) of Parental Education and Family Structure;*

Outcomes were regressed on the relevant exposure with each respondent assigned a weight equal to P(X)/P(X|C), where X represents respondents’ observed values for the exposure in question, and C represents a set of pre-exposure confounders as listed in Table 1 in the main paper. The purpose of this ATE weighting is to balance observed pre-exposure confounders (C) across levels of exposure (X). The probabilities required to calculate these weights were estimated via logistic regression models of X (with and without C).

Figure S1.3 shows standardised mean differences in pre-exposure confounders associated with low parental education, both before and after application of these ATE weights. While there were some clear observed differences in mother’s age at first live birth associated with low parental education, the ATE weighting reduced these differences to negligible levels. Figure S1.4 shows the impact of ATE weights for standardised mean differences associated with baseline lone parenting. While there were clear differences in parental education and mother’s age at first live birth associated with baseline lone parenting, the ATE weights reduced these to negligible levels.

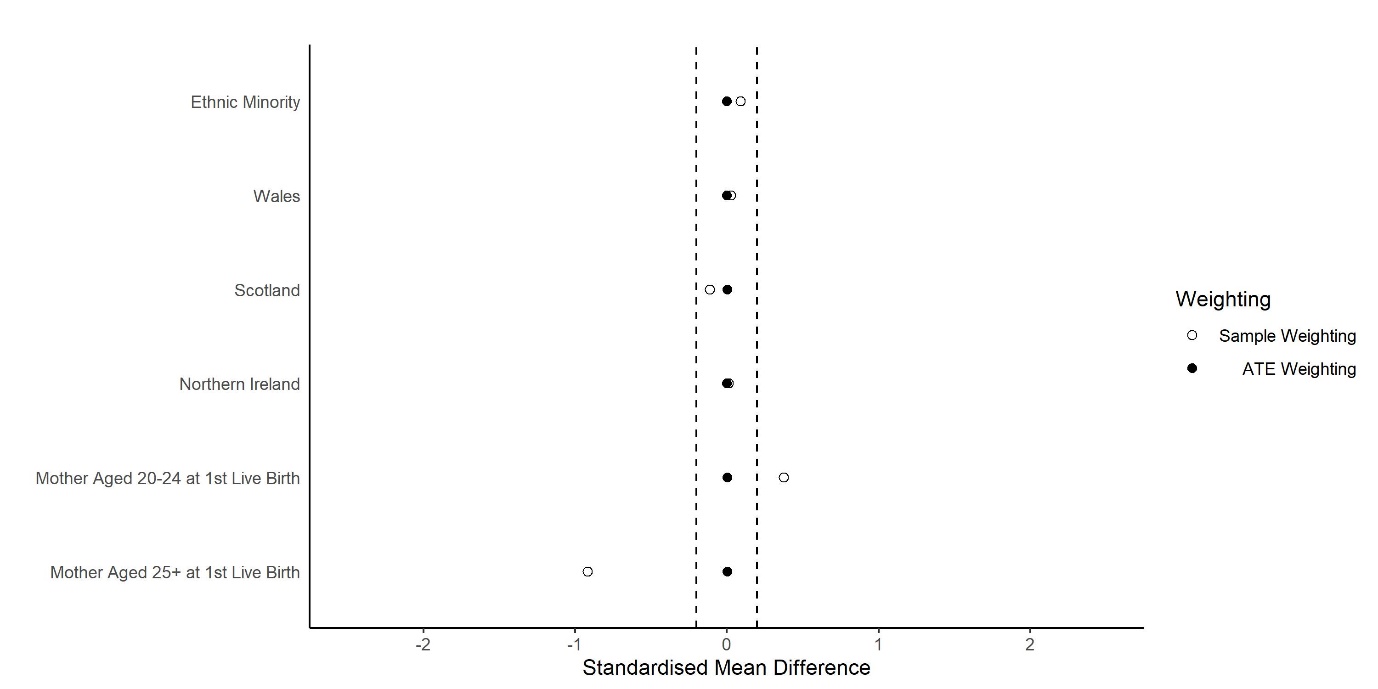

**Figure S1.3: Standardised Mean Differences in Confounders Associated with Low Parental Education**

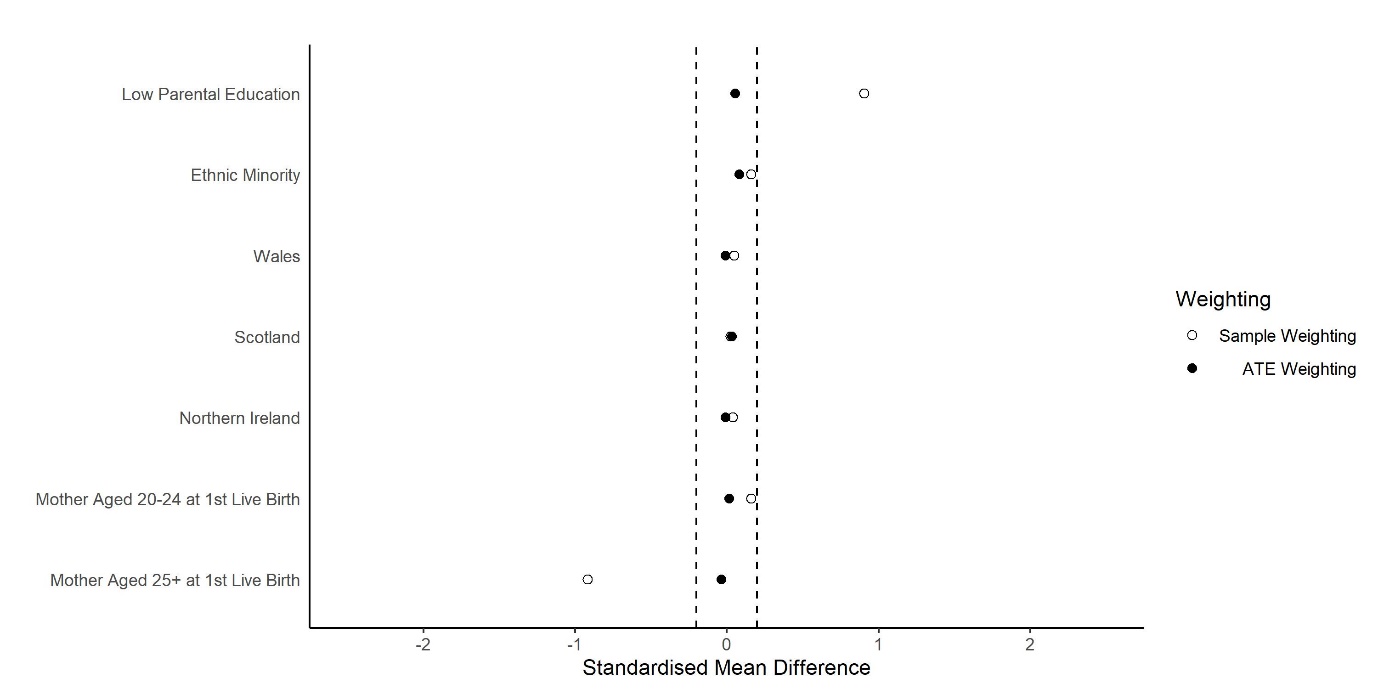

**Figure S1.4: Standardised Mean Differences in Confounders Associated with Baseline Lone Parenting**

*Estimating controlled direct effects:*

Outcomes were regressed on the exposure, mediator (childcare), and an interaction term for the two factors. These marginal structural models were weighted using a combination of the ATE weight for the exposure (as described above; P(X)/P(X|C)) and a modified version of the child-care weight, this time calculated as P(M|X)/P(M|X,C,L). As the numerator of this second weight is conditional on the exposure in question (X), it serves to balance post-exposure confounders (L), but only within levels of the exposure, so differences in these variables that are associated with the exposure are retained. Thus, performance of CDE weights can be assessed by examining standardised mean differences associated with childcare within levels of the exposure.

Standardised mean differences in confounders associated with childcare are shown in Figure S1.5 for those with high parental education and Figure S1.6 for those with low parental education. CDE weighting reduced all differences associated with centre-based (compared to parental) care within levels of parental education to negligible levels. The CDE weighting was less successful for non centre-based care and while it removed many of the observed differences (compared to parental care) within levels of parental education, some minor differences in confounders were still present, and may lead to some minor residual bias in the CDE estimates for parental education.

Figures S1.7 and S1.8 show standardised mean differences associated with childcare use for those with couple or lone parents at baseline respectively. The CDE weights were successful in removing differences in confounders associated with childcare use among baseline couple parents but were not successful in balancing these confounders among the smaller group of baseline lone parents (lone parents constituted 14.4% of the sample compared to 85.6% for couple parents). This may mean some residual bias is present in the CDE estimates for lone parenting, and suggests data were too sparse to fully disentangle differences in childcare among lone parents from differences in observed confounders. Nevertheless, despite these potential residual biases the considerable improvements in confounder balance among those with couple parents (the majority of the sample), mean that the CDE estimates for lone parenting probably still represent a net reduction in bias.

Figures S1.9-S1.12 repeat Figures S1.5-S1.8 for the alternate weights that adjusted for a more restricted set of confounding variables. Results were similar and suggested, if anything, slightly less residual bias than in the main analysis.

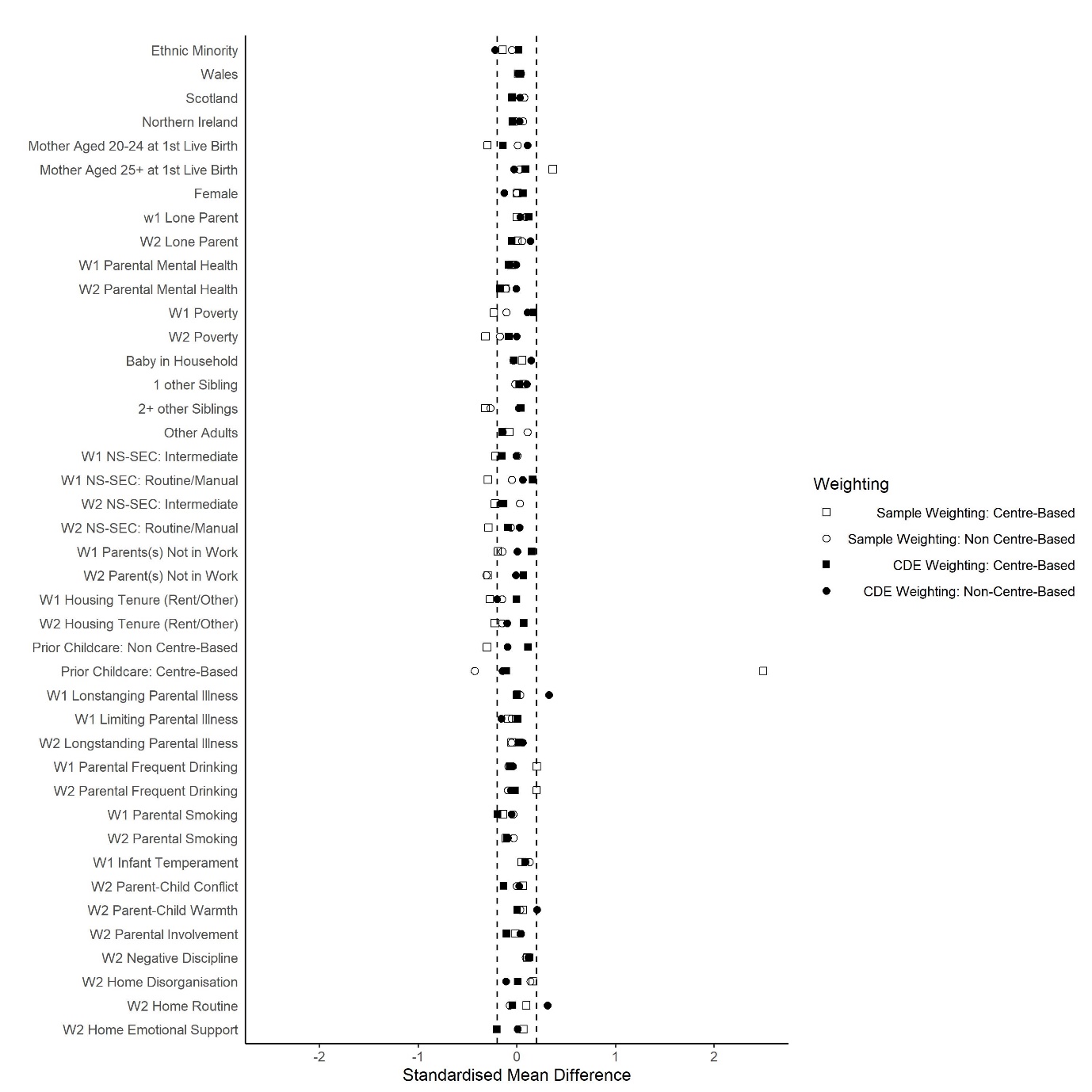

**Figure S1.5: Standardised Mean Differences in Confounders Associated with Childcare (High Parental Education)**

W1 indicates measures taken at 9 months, while W2 indicates measures taken at age 3.

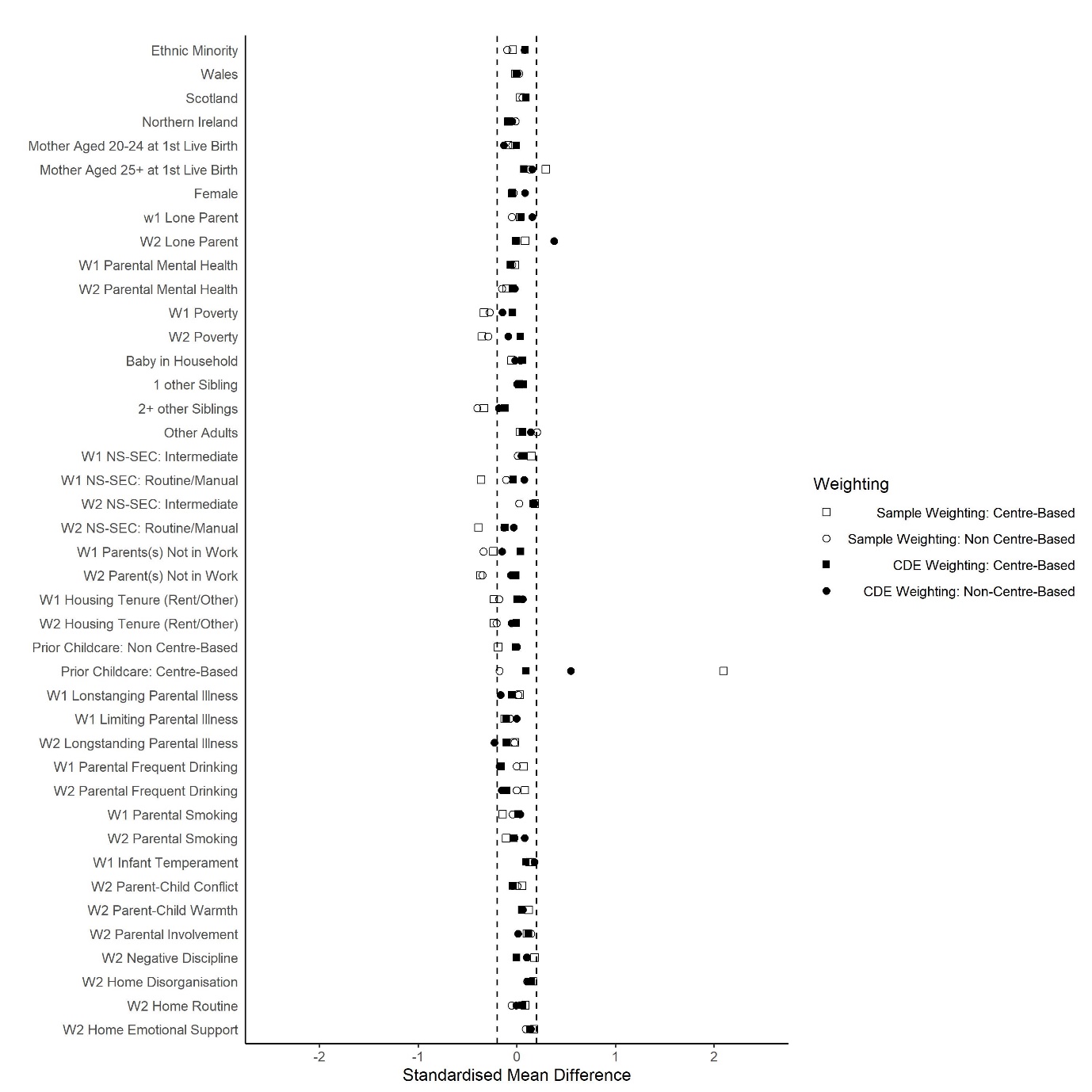

**Figure S1.6: Standardised Mean Differences in Confounders Associated with Childcare (Low Parental Education)**

W1 indicates measures taken at 9 months, while W2 indicates measures taken at age 3.

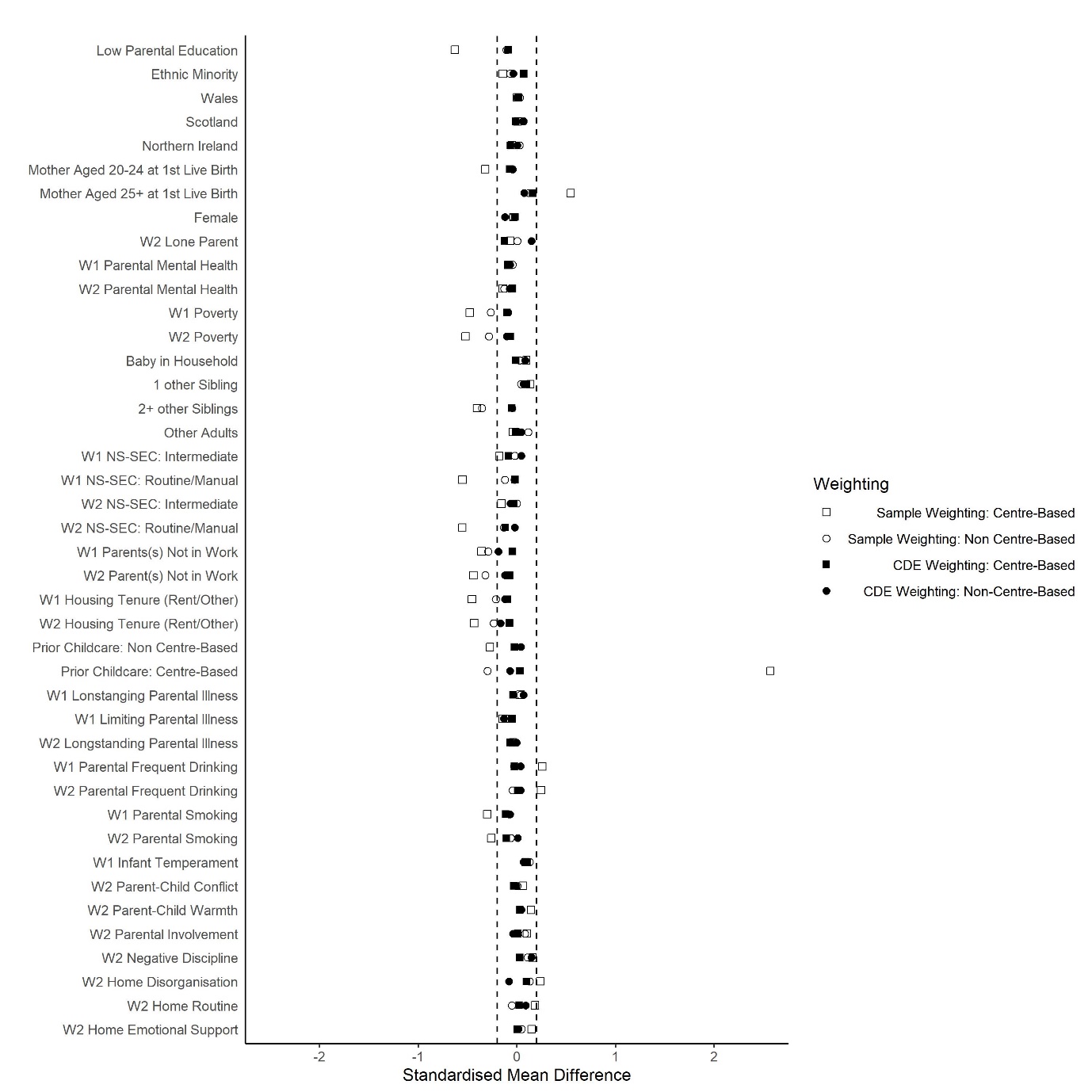

**Figure S1.7: Standardised Mean Differences in Confounders Associated with Childcare (Baseline Couple Parents)**

W1 indicates measures taken at 9 months, while W2 indicates measures taken at age 3.

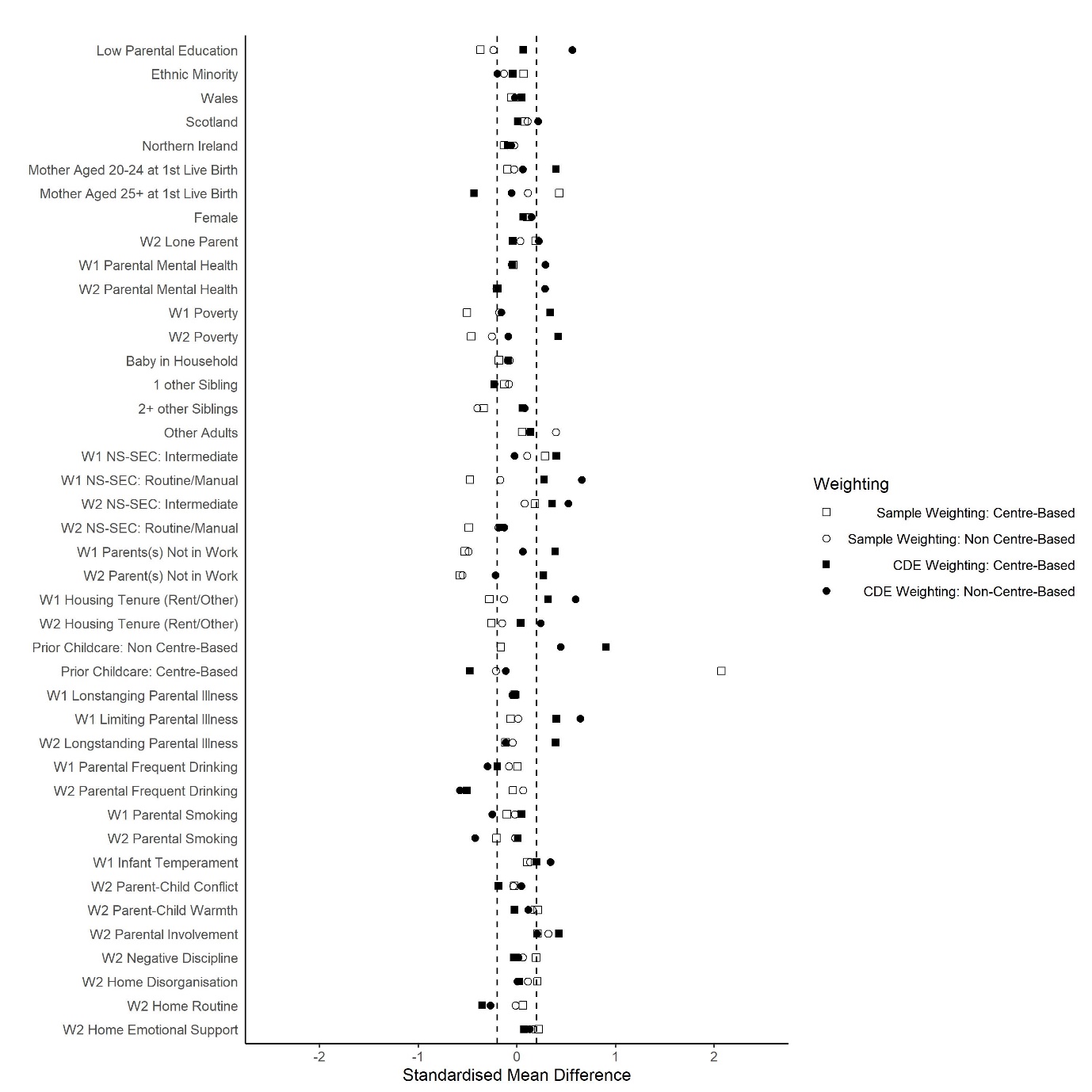

**Figure S1.8: Standardised Mean Differences in Confounders Associated with Childcare (Baseline Lone Parents)**

W1 indicates measures taken at 9 months, while W2 indicates measures taken at age 3.

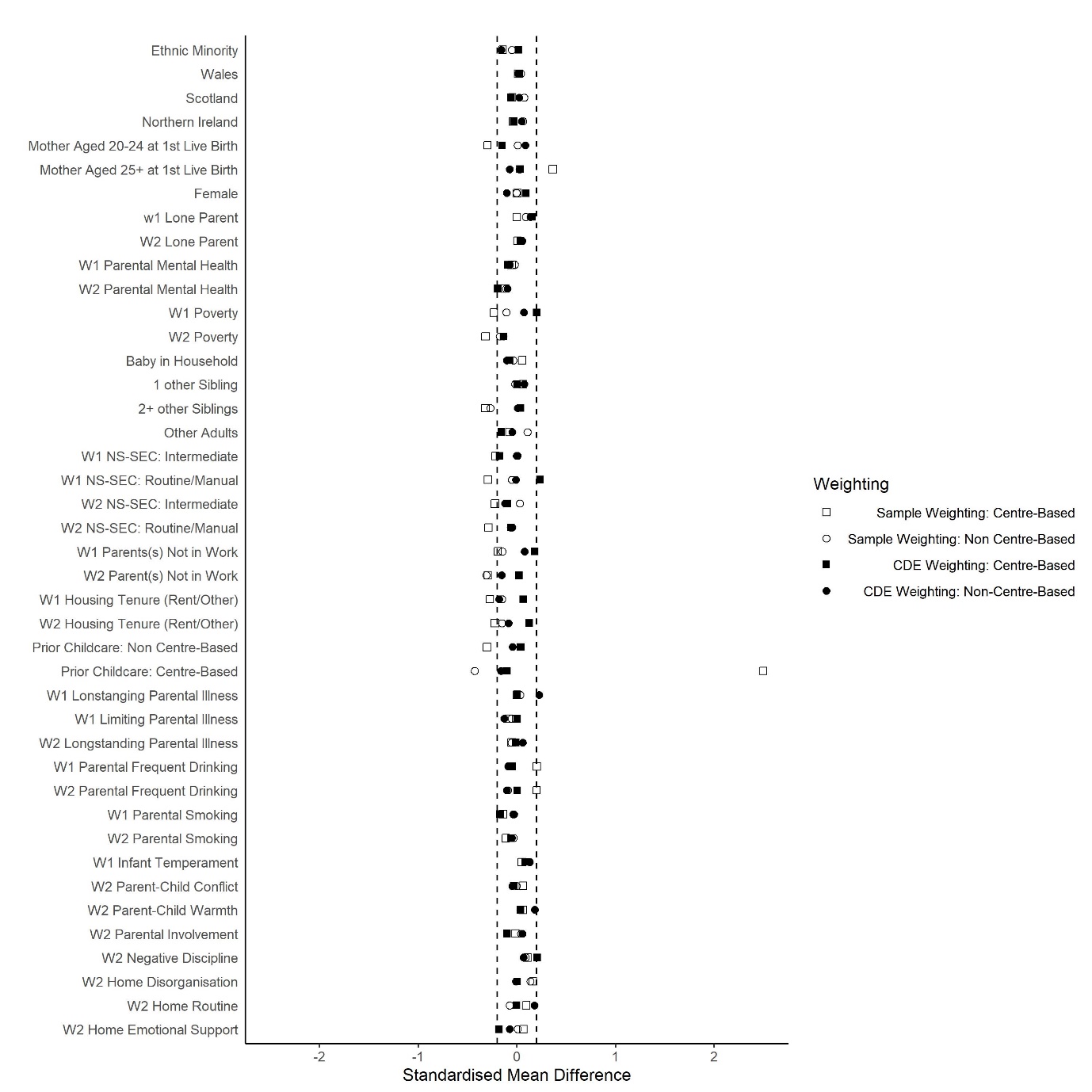

**Figure S1.9: Standardised Mean Differences in Confounders Associated with Childcare (High Parental Education; Alternate Weighting)**

W1 indicates measures taken at 9 months, while W2 indicates measures taken at age 3.

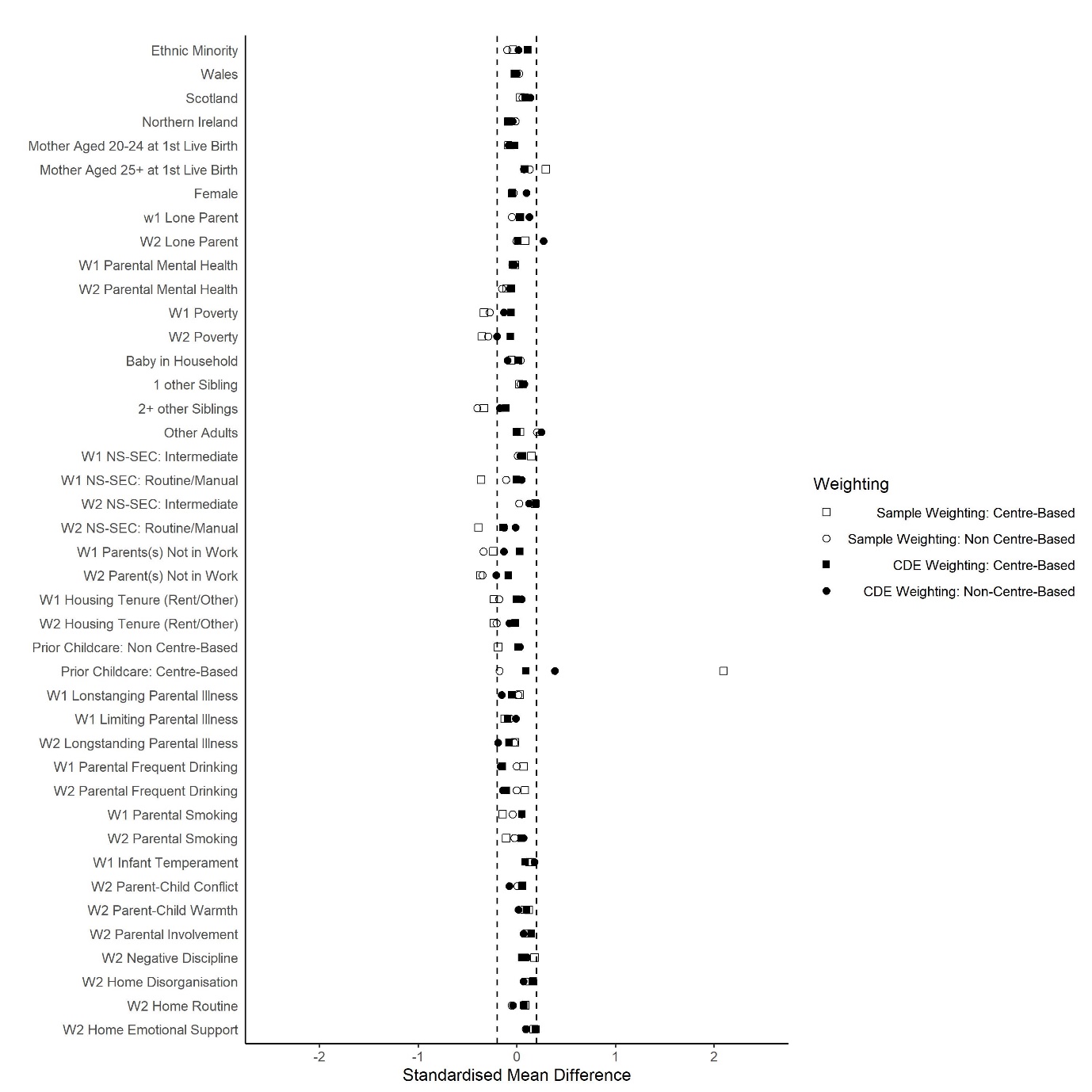

**Figure S1.10: Standardised Mean Differences in Confounders Associated with Childcare (Low Parental Education; Alternate Weighting)**

W1 indicates measures taken at 9 months, while W2 indicates measures taken at age 3.

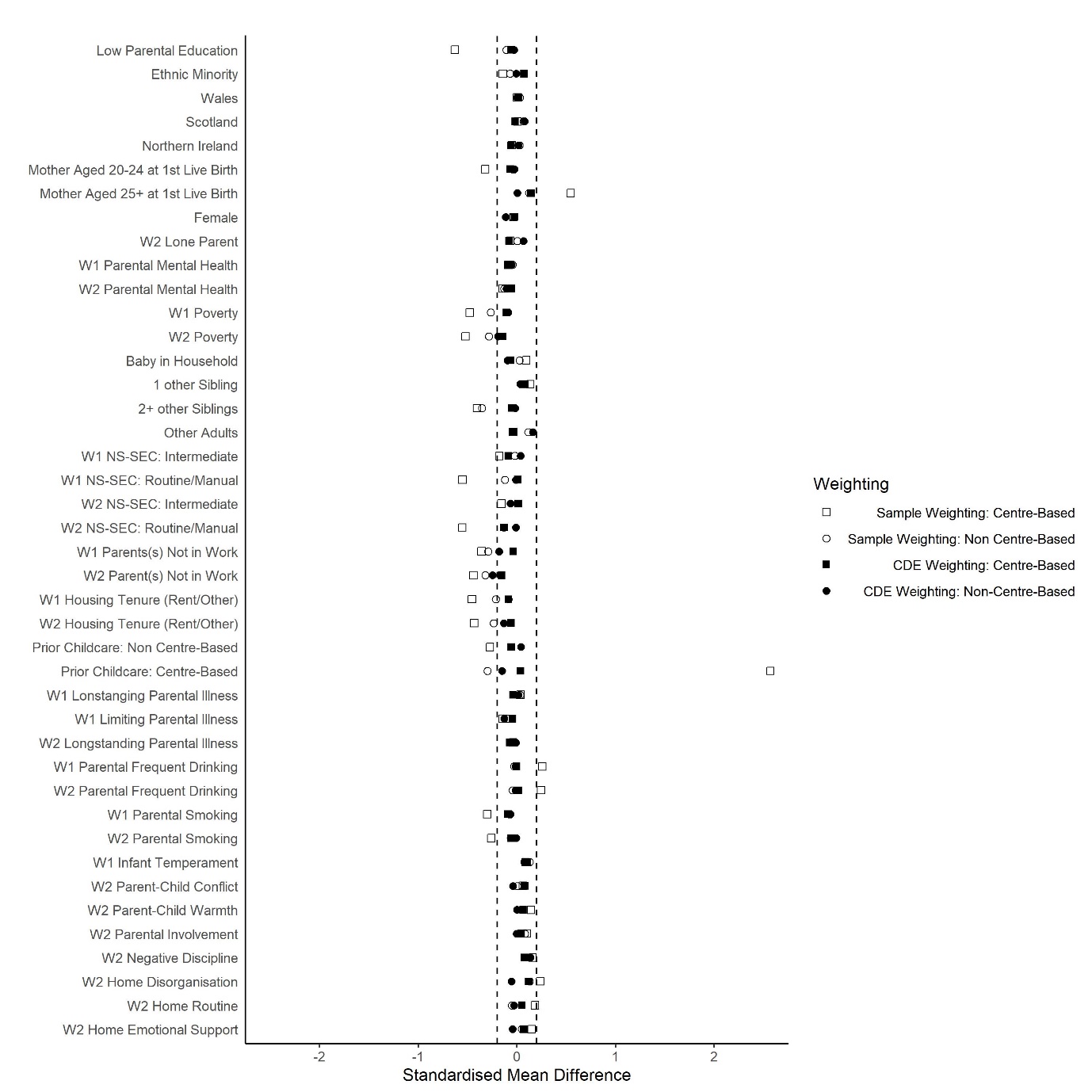

**Figure S1.11: Standardised Mean Differences in Confounders Associated with Childcare (Baseline Couple Parents; Alternate Weighting)**

W1 indicates measures taken at 9 months, while W2 indicates measures taken at age 3.

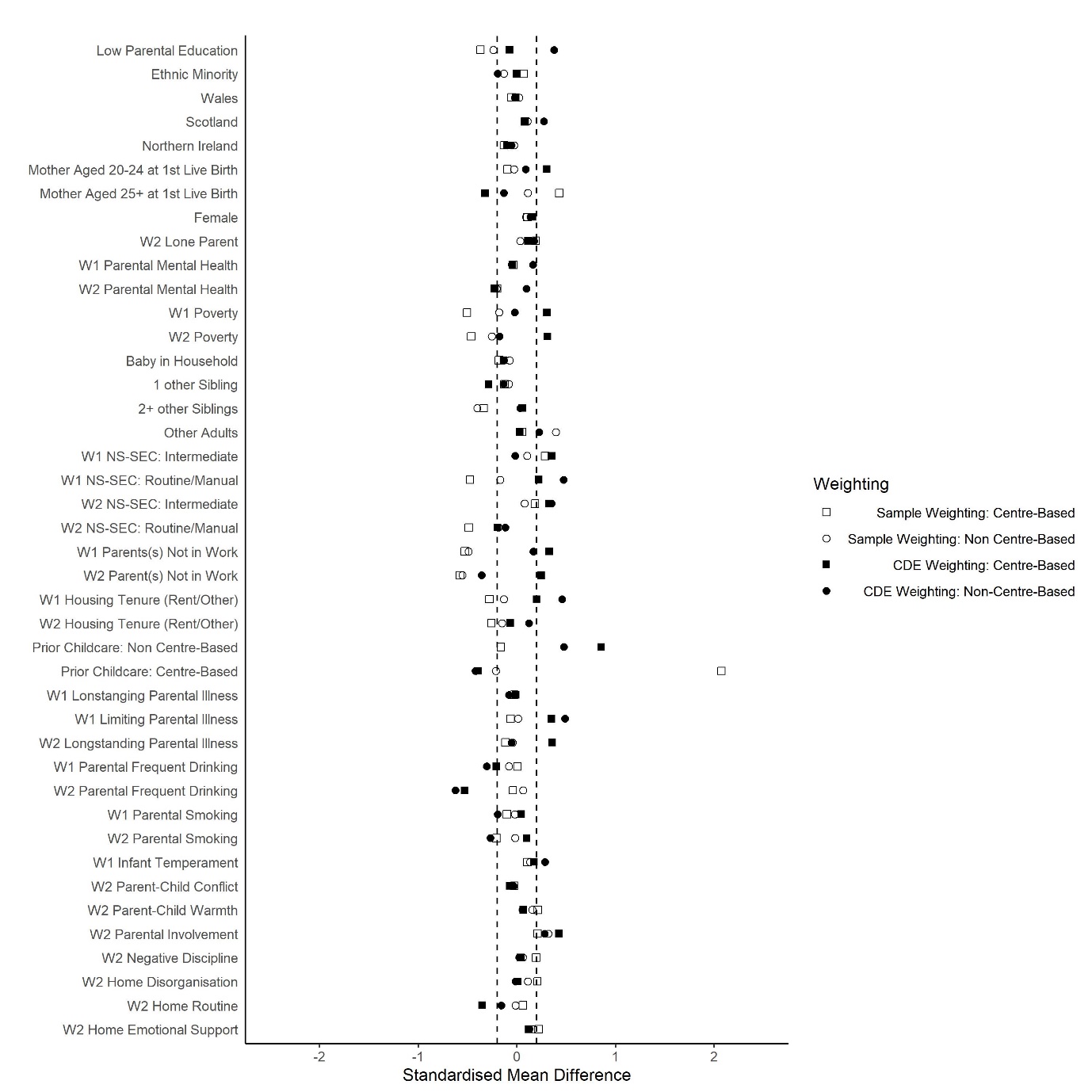

**Figure S1.12: Standardised Mean Differences in Confounders Associated with Childcare (Baseline Lone Parents; Alternate Weighting)**

W1 indicates measures taken at 9 months, while W2 indicates measures taken at age 3.
