## Supplementary File 2 for "Pre-school childcare and inequalities in child development"

Article Title: **Pre-school childcare and inequalities in child development**

Journal name: **European Journal of Epidemiology**

1. MRC/CSO Social & Public Health Sciences Unit, University of Glasgow.
2. Public Health Scotland.

**Supplementary File 2**

This supplementary file provides details of three sensitivity analyses. Each repeats the main analyses with a specific modification as follows:

1. Length of childcare exposure changed from 6 to 3 months.
2. Length of childcare exposure changed from 6 to 12 months.
3. Restricted set of post-exposure confounders.

*Sensitivity Analysis 1: 3 months’ childcare exposure*

In this analysis the childcare variable was altered to indicate exposure to childcare over a 3-month period (from 29-31 months of age). Figures S2.1-S2.3 are analogous to Figures 2-4 in the main paper, and show effect estimates of a similar magnitude.

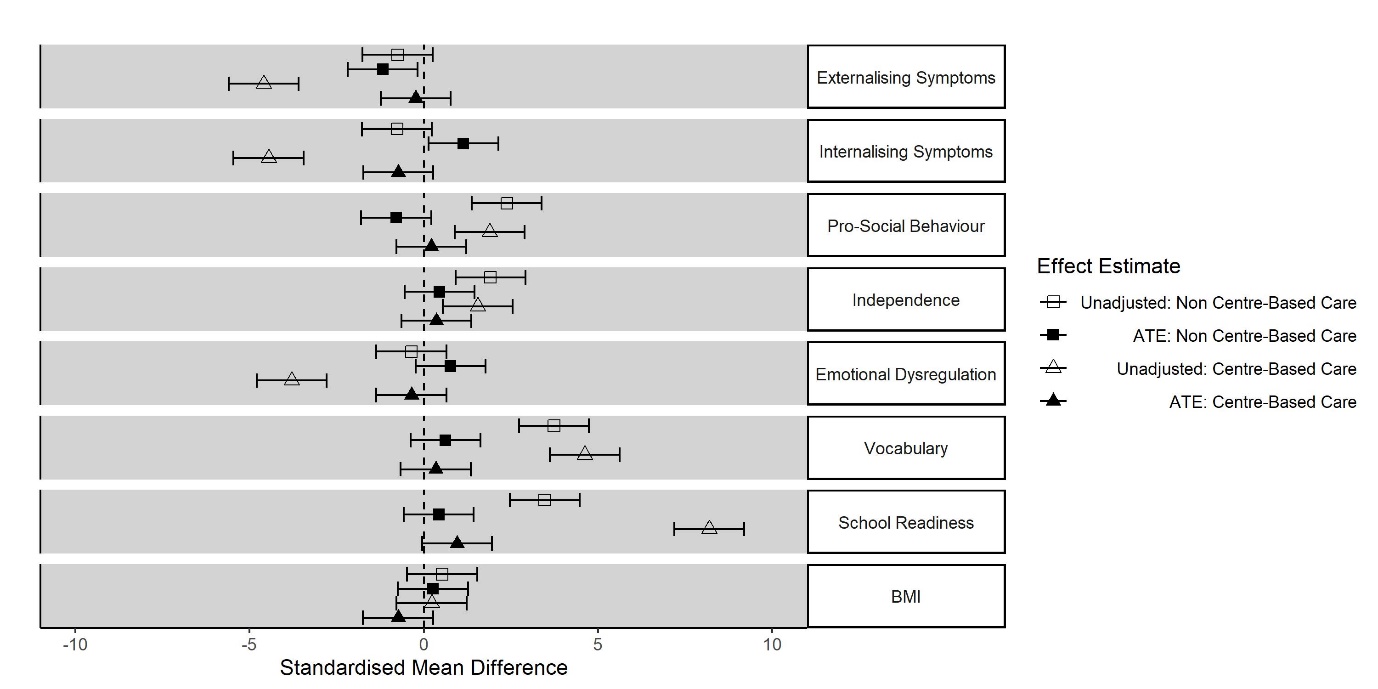

**Figure S2.1: Estimated effects of centre and non-centre-based childcare on child outcomes compared to parental care only (from ages 29-31 months)**

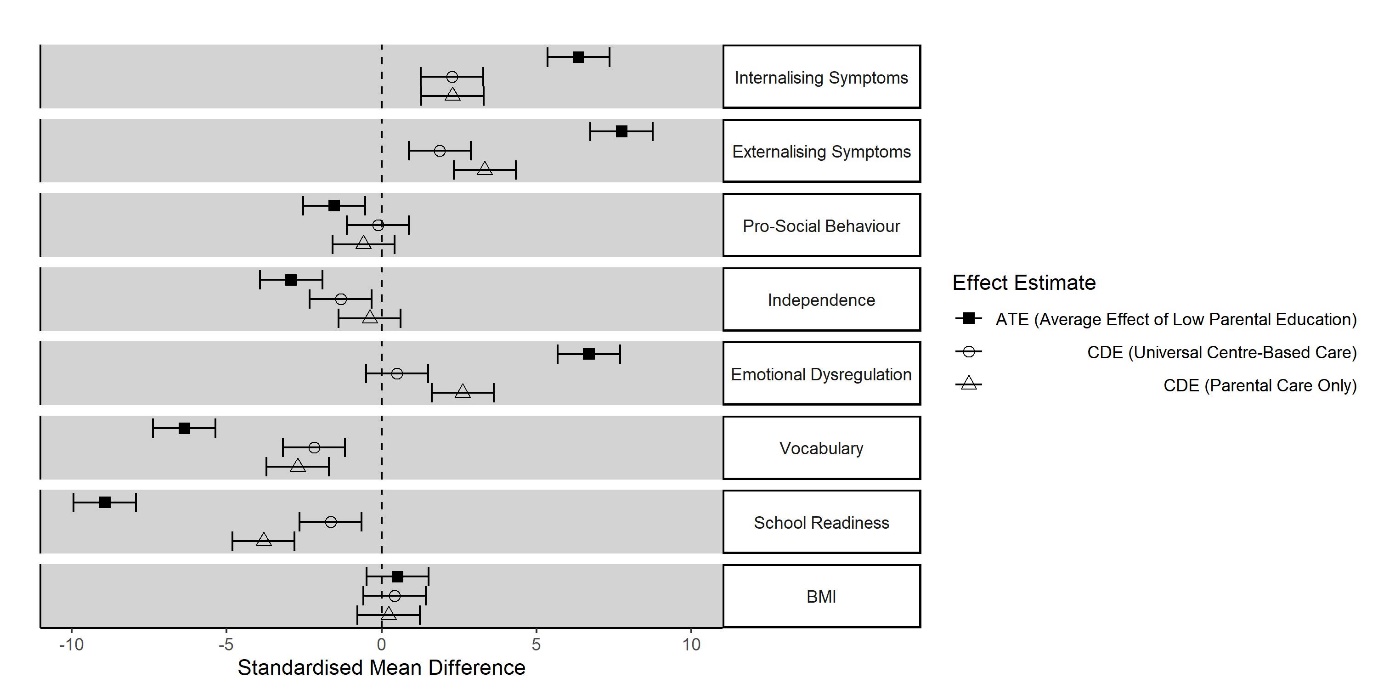

**Figure S2.2: Inequalities in child outcomes by parental education (Low vs. High), in the observed data (ATE), scenario 1 (universal centre-based care) and scenario 2 (parental care only)**

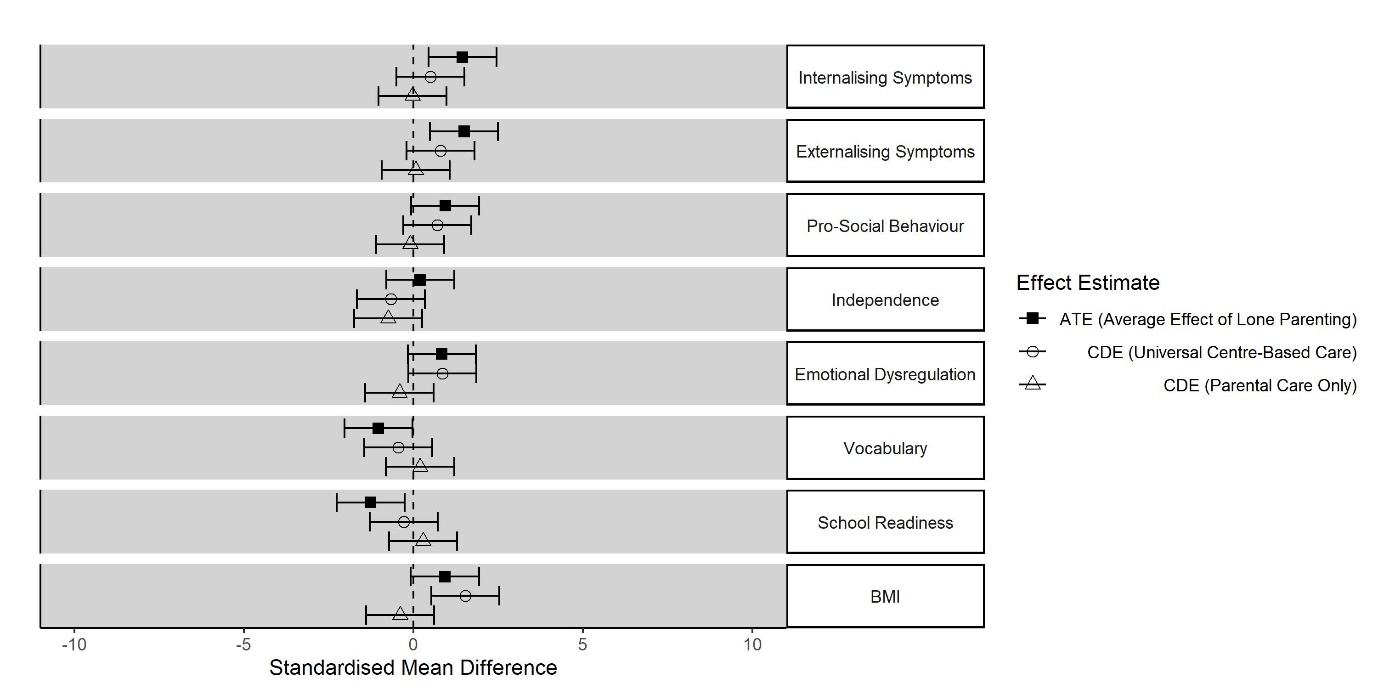

**Figure S2.3: Inequalities in child outcomes according to lone parenthood, in the observed data (ATE), scenario 1 (universal centre-based care) and scenario 2 (parental care only)**

*Sensitivity Analysis 2: 12 months’ childcare exposure*

In this analysis the childcare variable was altered to indicate exposure to childcare over a 12-month period (from 20-31 months of age). Figures S2.4-S2.6 are analogous to Figures 2-4 in the main paper, and show effect estimates of a similar magnitude.

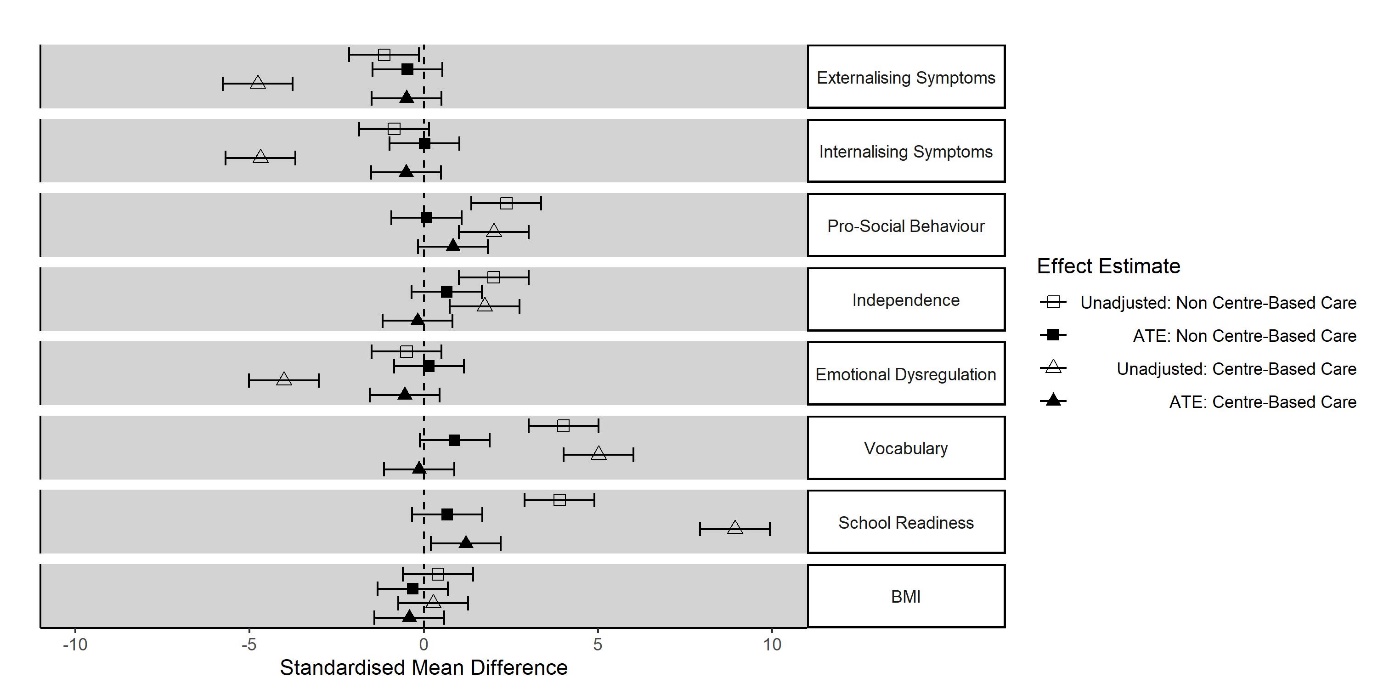

**Figure S2.4: Estimated effects of centre and non-centre-based childcare on child outcomes compared to parental care only (from ages 29-31 months)**

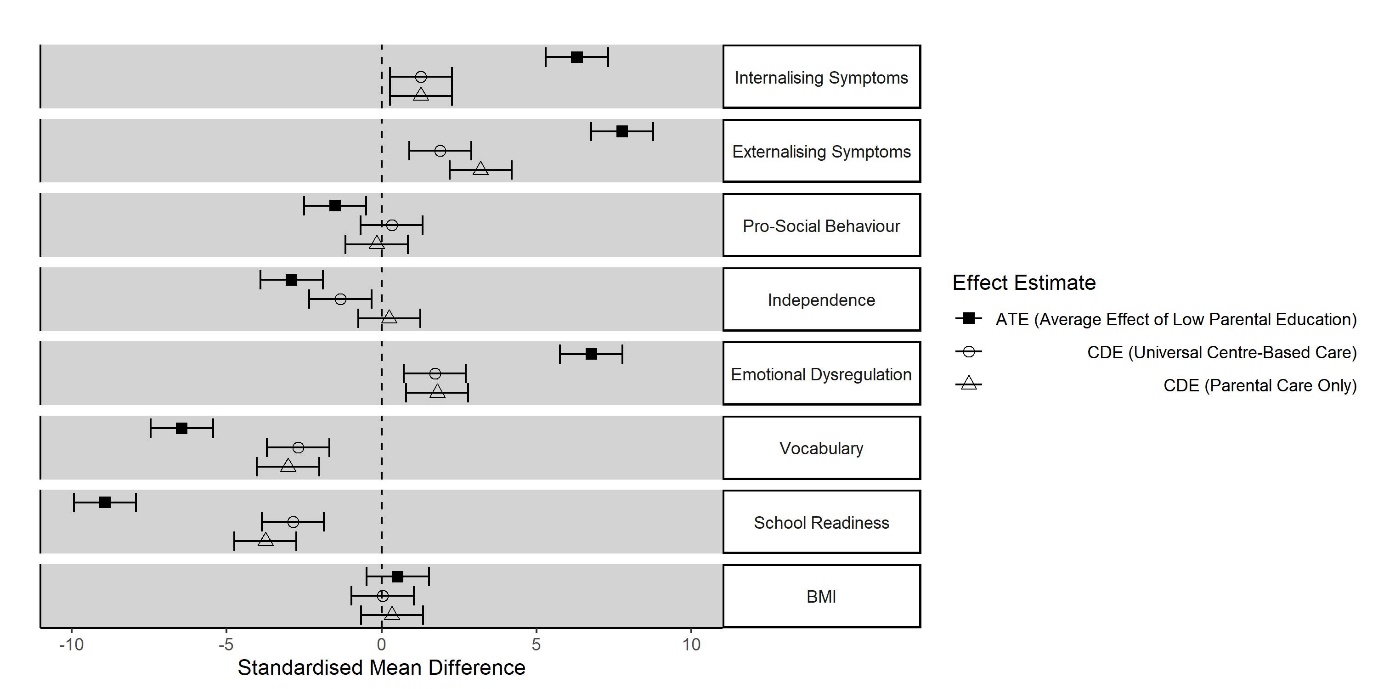

**Figure S2.5: Inequalities in child outcomes by parental education (Low vs. High), in the observed data (ATE), scenario 1 (universal centre-based care) and scenario 2 (parental care only)**

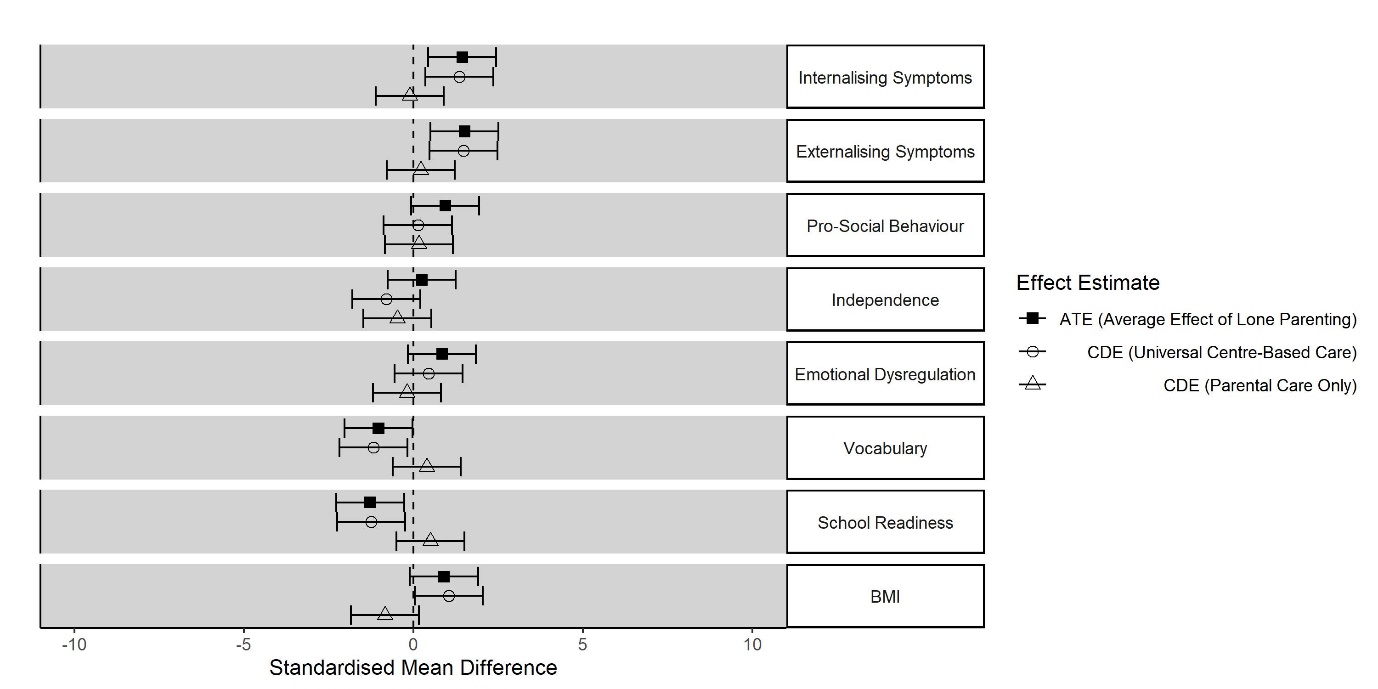

**Figure S2.6: Inequalities in child outcomes according to lone parenthood, in the observed data (ATE), scenario 1 (universal centre-based care) and scenario 2 (parental care only)**

*Sensitivity Analysis 3: Restricted Confounder Set*

In this analysis a restricted set of post-exposure confounders (L) was included on the basis that some of these variables may have been determined by rather than being determinants of childcare. Figures S2.7-S2.9 are analogous to Figures 2-4 in the main paper, and show effect estimates of a similar magnitude.

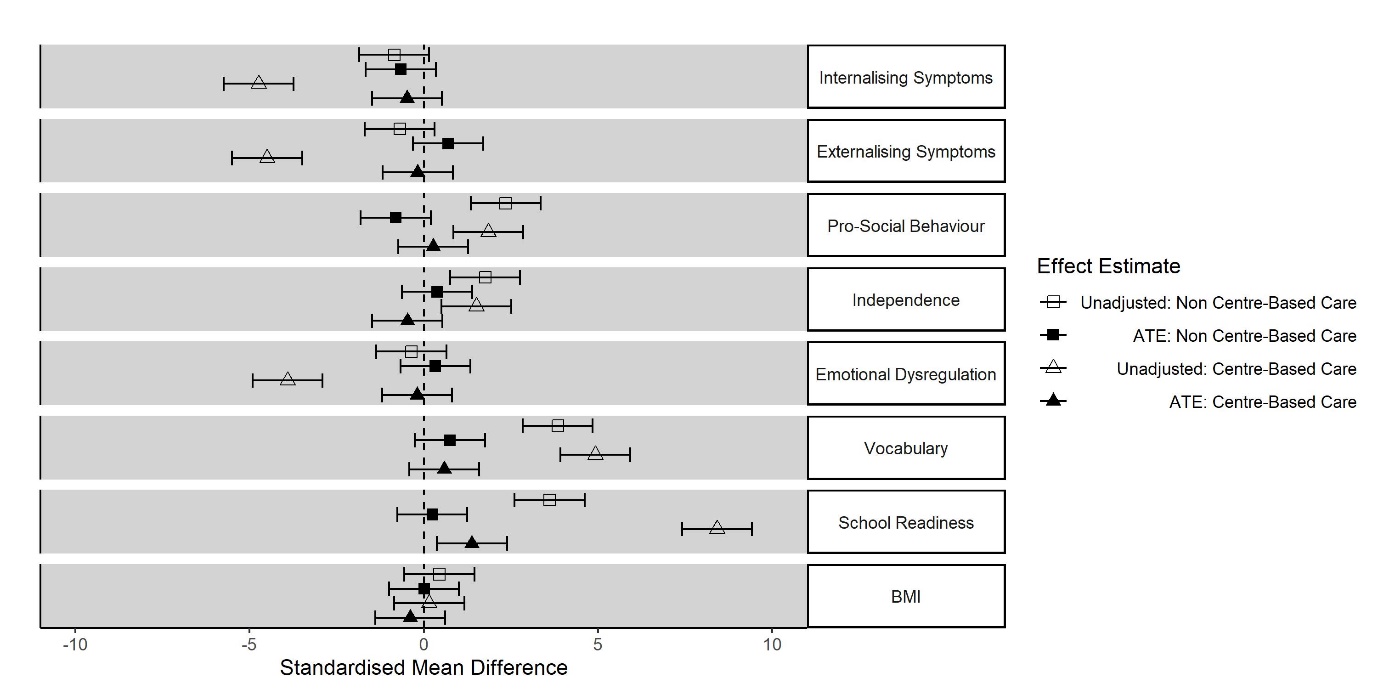

**Figure S2.7: Estimated effects of centre and non-centre-based childcare on child outcomes compared to parental care only (from ages 26-31 months) -restricted confounder set**

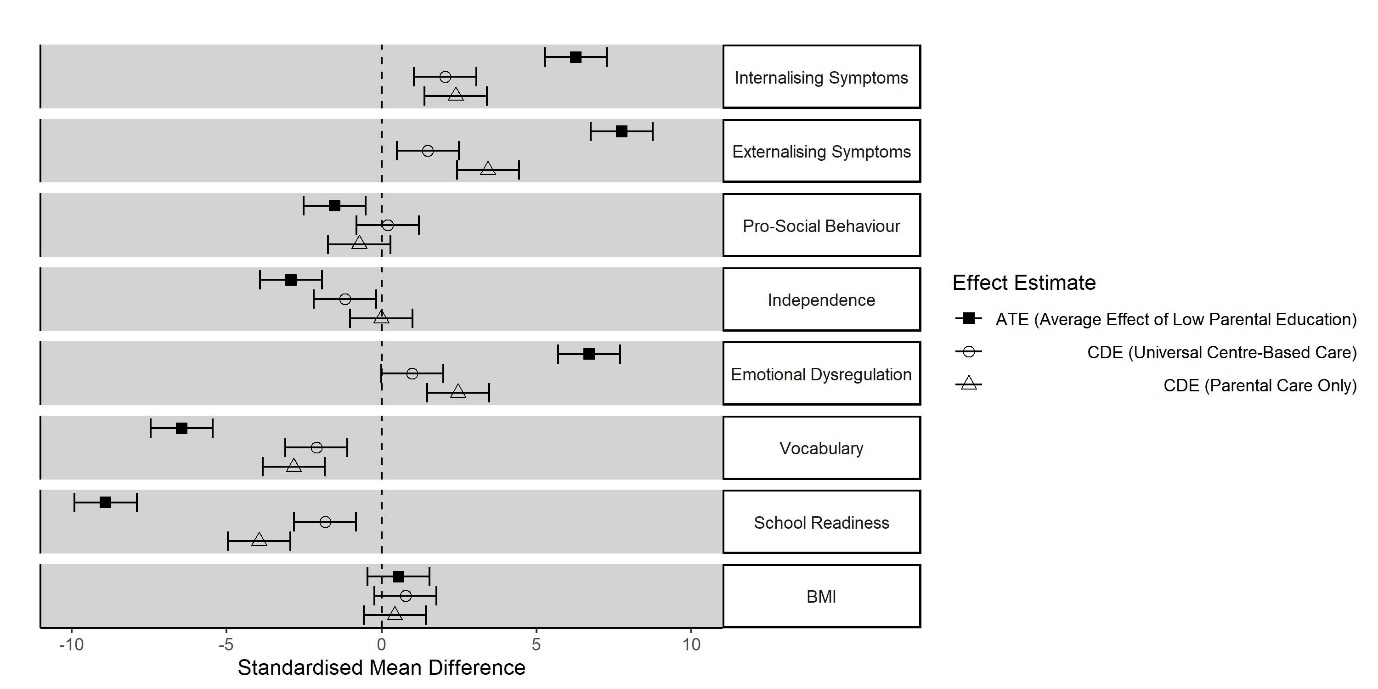

**Figure S2.8: Inequalities in child outcomes by parental education (Low vs. High), in the observed data (ATE), scenario 1 (universal centre-based care) and scenario 2 (parental care only) -restricted confounder set**

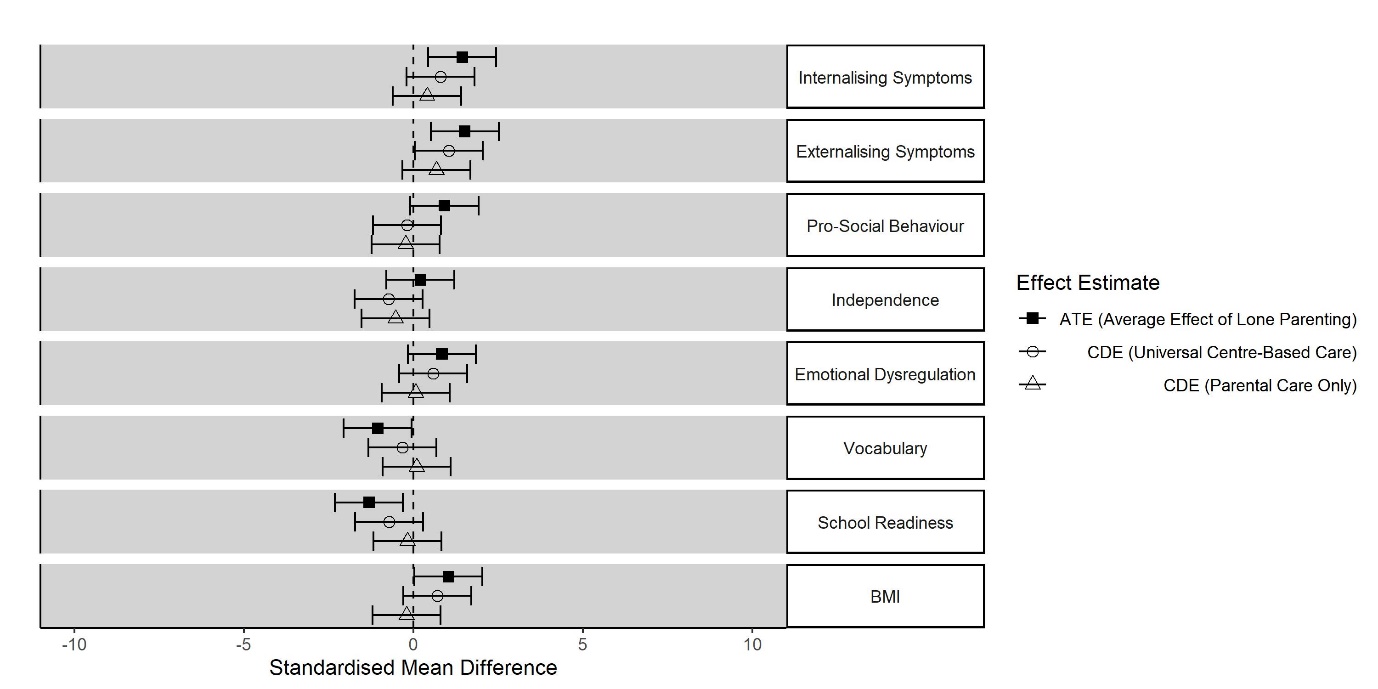

**Figure S2.9: Inequalities in child outcomes according to lone parenthood, in the observed data (ATE), scenario 1 (universal centre-based care) and scenario 2 (parental care only) -restricted confounder set**
